# Combining MAVEs and computational predictors improves variant classification across ancestries in hereditary cancer genes

**DOI:** 10.64898/2025.12.08.25341119

**Authors:** Nikita Bora, Mihaly Badonyi, Ankit K Pathak, Rutharra Ghayadthri Manisekaran, SG10K_Health Consortium, Joseph A Marsh, Joanne Ngeow

**Affiliations:** Lee Kong Chian School of Medicine, Nanyang Technological University, Singapore; MRC Human Genetics Unit, Institute of Genetics and Cancer, University of Edinburgh, Edinburgh, UK; Cancer Genetics Service, Division of Medical Oncology, National Cancer Centre Singapore, Singapore

**Author notes:** These authors have contributed equally to this work (Nikita Bora, Mihaly Badonyi, Ankit K Pathak). A complete list of contributing authors to the SG10K_Health Consortium is provided in Table S1.

## Abstract

Many commonly used computational tools for variant effect prediction exhibit ancestry-related bias because they are trained on clinical or population datasets that under-represent global diversity, leading to uneven and sometimes unfair variant classification across ancestries. Multiplexed assays of variant effect (MAVEs) and population-free VEPs instead offer alternatives that are unbiased with respect to human ancestry, providing classification evidence that generalises across populations. Here, we evaluate MAVE- and VEP-based classification across five cancer-associated genes with high-quality MAVE datasets, focusing on three leading population-free VEPs: GEMME, EVE, and CPT-1. We find that MAVEs are more conservative and decisive in classification, assigning fewer variants to the pathogenic category while yielding fewer indeterminate classifications when calibrated to ACMG/AMP guidelines. While MAVEs show heightened sensitivity in functionally assayed regions, VEPs identify a broader range of pathogenic variants overall. By combining clinical evidence strengths from MAVEs and VEPs, we reclassify over 90% of variants of uncertain significance across the SG10K_Health and Mexico City Prospective Study reference datasets as at least likely benign or likely pathogenic. We further isolate variants where MAVE- and VEP-based classifications are discordant, highlighting method-specific limitations. Together, these findings clarify the complementary strengths of experimental and computational classification approaches and provide a path to less biased and more equitable variant interpretation in clinical genomics, helping to mitigate disparities in diagnosis across ancestries.

## Introduction

High-throughput sequencing now delivers vast catalogues of human genetic variation, but our ability to interpret that variation has not kept pace. A substantial proportion of variants detected in clinical testing remain classified as of uncertain significance, limiting their diagnostic value and complicating patient management [1,2]. This interpretive gap reflects the diversity of molecular mechanisms by which variants act and the difficulty of distinguishing deleterious from tolerated changes at scale. Addressing it requires scalable approaches that provide reliable and clinically meaningful evidence of variant impact.

Computational variant effect predictors (VEPs) have become indispensable for this task, with their outputs now carrying growing weight in clinical variant interpretation frameworks such as the ACMG/AMP guidelines [3–5]. Yet, despite their widespread use, many VEPs are subject to biases inherited from their training data. Predictors trained on patient-derived variants can overestimate performance when benchmarked on similar datasets, obscuring generalisability [6,7]. Other VEPs incorporate population-derived variants such as allele frequencies and may therefore be affected by ancestry-related biases, as current sequencing data remains heavily skewed towards individuals of European descent, with significant implications for public health equity [8–10]. Crucially, several population-free VEPs, trained without clinical or population variant labels, now match or exceed top-performing predictors [7] while minimising any risk of circularity and avoiding the possibility of ancestry-related bias [9].

In parallel with the development of computational approaches, multiplexed assays of variant effect (MAVEs) have become a powerful experimental platform for the systematic interrogation of the functional effect of missense variants [11,12]. By combining saturation mutagenesis with high-throughput functional assays and deep sequencing, MAVEs can infer variant effects on protein activity, abundance or the growth rate of cells [13–15]. Compared to computational predictions from VEPs, MAVEs potentially provide more direct representations of variant impacts, particularly when the experimental setup closely mimics physiologically and clinically relevant conditions.

Thus, MAVEs and population-free VEPs have emerged as promising alternatives for variant classification that are unbiased with respect to ancestry, but their relative strengths remain poorly understood. MAVEs are often limited to specific genes or assay conditions, whereas population-free VEPs, although applicable across the proteome, may fail to capture subtle or context-dependent effects. These differences raise important questions about how the classification properties of the two methods compare, particularly in clinical variant prioritisation and triage.

To address this, we systematically compared MAVE- and VEP-based classification across five hereditary cancer genes with high-quality functional datasets: *BRCA1* [16], *HRAS* [17], *MSH2* [18], *PTEN* [19], and *TP53* [20]. Within each gene, we compared calibrated MAVE and VEP score distributions to identify intermediate-impact variants, mapped concordant and discordant calls onto protein structures and ΔΔG-based stability metrics to expose mechanistic patterns, and converted calibrated scores into ACMG/AMP evidence strengths to assess their combined utility for reclassification. Finally, to test the translational value of these approaches, we applied them to population-scale variant data from the Singapore SG10K_Health cohort [21,22] and the Mexico City Prospective Study [23]. These datasets allow us to evaluate classification in ancestries under-represented in existing clinical and population references and to assess whether integrating MAVEs with population-free VEPs can resolve variants of uncertain significance robustly across global populations. By examining their relative strengths and weaknesses, we show how this integration can promote more equitable and clinically meaningful evaluation and inform their future use in variant interpretation.

## Results and discussion

### Performance of MAVEs and VEPs on five cancer genes

We assembled missense variant datasets for five genes primarily linked to cancer, although their mutations are not exclusively cancer-related. To ensure comparability, we limited our analysis to variants that overlap with those surveyed by the corresponding MAVE assays. For *BRCA1*, *MSH2*, and *TP53*, we classified ClinVar [24] benign or likely benign (B/LB) variants as ‘benign’. For *PTEN* and *HRAS*, two highly constrained dominant genes with insufficient numbers of ClinVar B/LB variants, we instead used variants observed in gnomAD [25], which we consider ‘putatively benign’. We note that while the gnomAD sets may include some disease-associated variants, the vast majority are expected to be functionally neutral in these genes, especially given the autosomal dominant nature of the known pathogenic variants. Pathogenic and likely pathogenic (P/LP), hereafter ‘pathogenic’, variants were drawn exclusively from ClinVar. A detailed breakdown of the number of variants surveyed by the MAVE assays, along with their overlaps with pathogenic and (putatively) benign variants, is summarised in **Table 1**.

**Table 1.** Summary of missense variant counts and details of the MAVE assays. An asterisk (*) marks putatively benign variants from gnomAD.

| Gene | Variants | Pathogenic | (Putatively*) benign | Reference |
| --- | --- | --- | --- | --- |
| *BRCA1* | 1,837 | 159 | 82 | Findlay et al., 2018 |
| *HRAS* | 3,135 | 29 | 182* | Bandaru et al., 2017 |
| *MSH2* | 16,749 | 128 | 243 | Jia et al., 2021 |
| *PTEN* | 6,564 | 195 | 190* | Mighell et al., 2018 |
| *TP53* | 7,448 | 195 | 122 | Giacomelli et al., 2018 |

We collected variant effect scores from three population-free VEPs, GEMME [26], EVE [27], and CPT-1 [28], chosen for their top performance in recent benchmarking [7]. GEMME is an empirical method that predicts the effects of mutations by modelling the evolutionary history of related sequences from a multiple sequence alignment. EVE is another empirical method that employs a variational autoencoder to learn the rules of an alignment and score variants accordingly. Lastly, CPT-1 is a cross-protein transfer model integrating two population-free VEPs, EVE and the language model ESM-1v [29], along with features extracted from predicted protein structures. While CPT-1 was additionally trained on MAVE data, this does not include any of the five genes that are the focus of this study.

We assessed the performance of MAVE and VEP scores in discriminating pathogenic and benign/putatively benign variants by calculating the area under the receiver operating characteristic (AUROC) curve. The results are visualised in **Fig. 1** as histograms, showing binned variant counts separately for the two classes and for each gene and predictor.

**Fig. 1.**
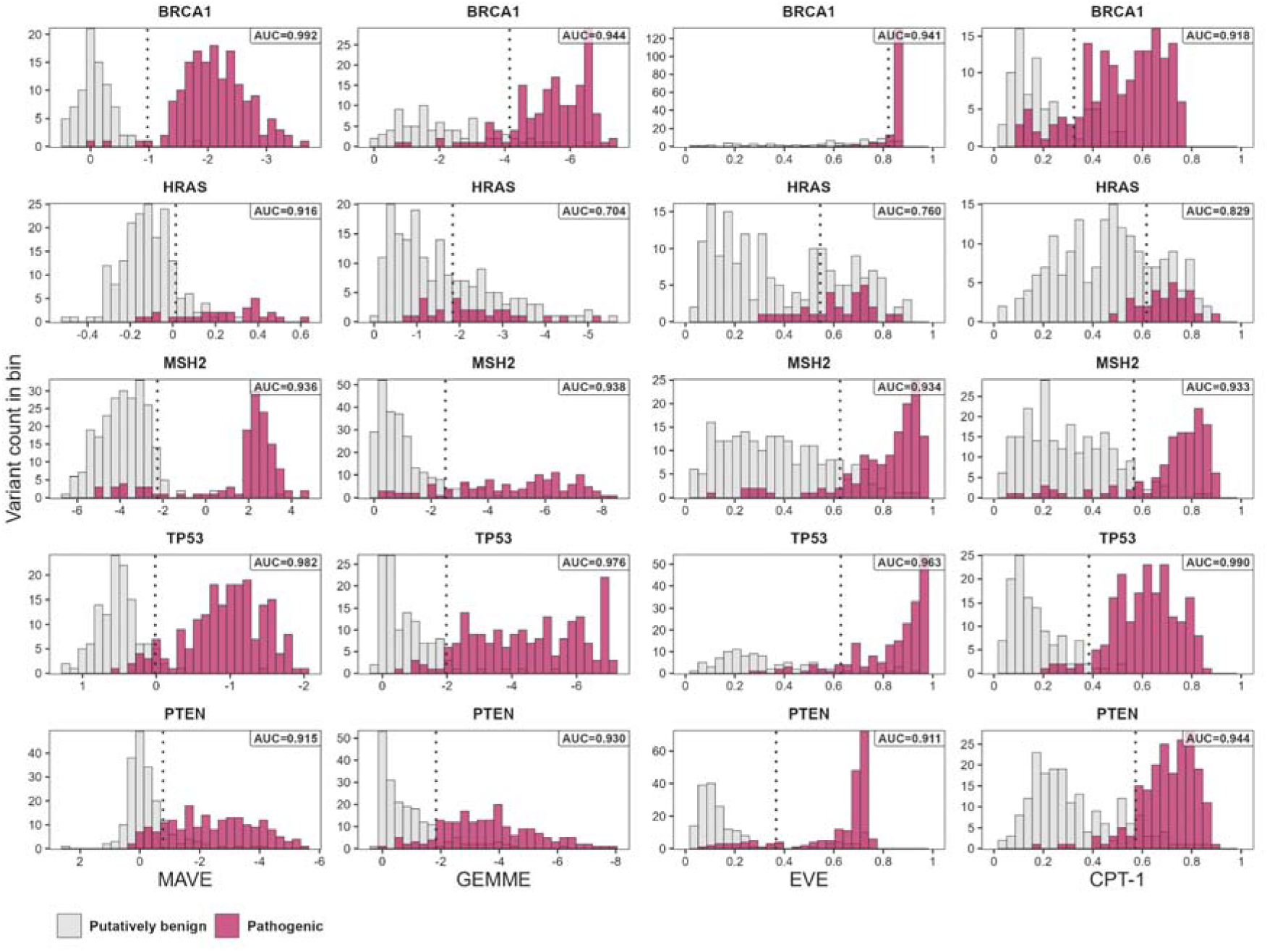
MAVE and VEP score distribution and performance. Histograms show the binned (N = 30) MAVE and VEP scores across the five genes, with the distribution of missense putatively benign variants from gnomAD (HRAS and PTEN) or missense benign variants from ClinVar (*BRCA1*, *MSH2*, and *TP53*) and missense pathogenic variants from ClinVar highlighted in grey and pink, respectively. Values in the top right corner represent the areas under the receiver operating characteristic (ROC) curves. Dotted vertical lines show the optimal score threshold, that is, the distance from the (0,1) point of the ROC curve, separating the two classes. The *x*-axes for GEMME and MAVEs of *BRCA1*, *PTEN,* and *TP53* are reversed such that pathogenic variants will tend to fall on the right side of the distribution.

MAVEs outperformed all three VEPs in the BRCA1 and HRAS assays. In contrast, GEMME and CPT-1 showed slightly better performance by AUROC than MAVEs in MSH2 and TP53, respectively, and both VEPs outperformed the functional assay in PTEN. Overall, CPT-1 emerged as the best-performing VEP in three of the five genes, consistent with a recent benchmarking study where it ranked first out of 97 tested VEPs [7].

The increased performance of MAVEs in BRCA1 and HRAS may be attributed to specific characteristics of these genes and the assay conditions. The essential role of BRCA1 in human HAP1 cells employed in the experiment likely enhances the sensitivity of the assay [16], making it particularly effective at detecting deleterious loss-of-function mutations. For *HRAS*, the presence of GTPase-activating proteins (GAP) in the assay without guanine nucleotide exchange factors (GEF) unmasks gain-of-function mutations [17] that evolution-based VEPs may currently be unable to detect from an alignment alone [30,31], giving MAVE a distinct advantage. Conversely, the MAVE assay for PTEN, which uses a humanised yeast system to measure intrinsic lipid phosphatase activity, may miss mutations that disrupt protein-protein interactions, subcellular localisation, post-translational modifications, or those exerting a dominant-negative effect in humans [19,32]. This limitation explains why all three VEPs outperform MAVE data in PTEN, as they may capture a broader spectrum of functional impacts.

### MAVE-aided stratification is more conservative yet broadly consistent with VEPs

Although binary pathogenic versus benign classifications are the standard for clinical interpretation, many missense variants have effects that are neither fully neutral nor strongly deleterious. Such intermediate-impact variants may contribute to disease through reduced penetrance, modifier effects, or partial loss of function, yet they are difficult to capture with conventional predictors. MAVEs, by directly assaying variant function in relevant biological contexts, may be particularly sensitive to these subtle effects, whereas VEPs, which rely primarily on evolutionary conservation and global sequence patterns, are expected to prioritise only the most deleterious substitutions. To explore this distinction, we extended our framework to include an intermediate classification category and compared how MAVE- and VEP-based stratification capture these variants.

We classified variants into *mild*, *intermediate*, and *damaging* categories using MAVE and VEP scores based on the following criteria. Variants covered by the fitness maps with values above the optimal threshold separating pathogenic from benign/putatively benign variants were classified as damaging. (Note that in some cases, where higher scores indicate benign variants, the pathogenic threshold corresponds to lower scores.) The remaining variants were classified as mild, except for the 25% closest to the pathogenic threshold, which were classified as intermediate. A schematic representation of this classification procedure is shown in **Fig. 2A**. Counts of variants assigned to each class for the five genes are shown in **Fig. 2B**. As this framework is not intended for clinical classification, we refer to it hereafter as ‘MAVE- or VEP-aided stratification’ instead.

**Fig. 2.**
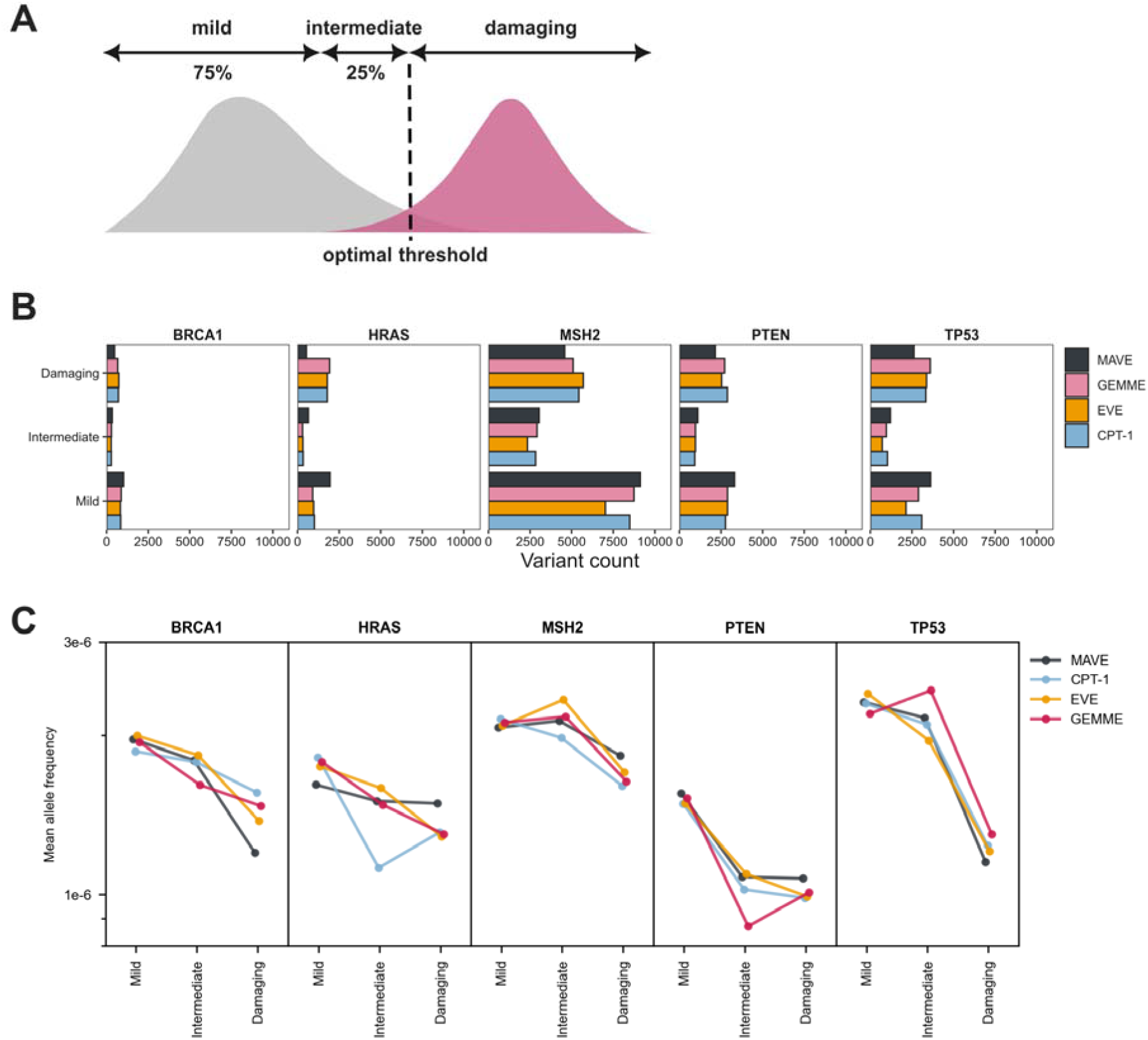
Variant counts and allele frequency distributions with MAVE- and VEP-aided stratification. **A:** Schematic representation of the MAVE- and VEP-aided stratification approach. Given the score distributions of pathogenic and (putatively) benign variants, an optimal threshold is derived to separate the two classes. Variants scoring above this threshold (or below, for inverted predictors) are classified as damaging. Among variants below the threshold, the bottom 75% are considered mild, while the top 25% are classified as intermediate-effect. **B**: Variant counts resulting from MAVE- and VEP-aided stratification. **C**: Distribution of average gnomAD v4.1 allele frequencies across mild, intermediate, and damaging classes.

We first examined the relationship between variant stratifications and their allelic prevalence in human populations (**Fig. 2C**). We expected mild, intermediate, and damaging variants to follow a gradient reflecting the effects of purifying selection: mild variants should exhibit the highest average allele frequencies, followed by intermediate variants, with damaging variants showing the lowest due to stronger selection. Although factors such as genetic drift, founder effects, and population bottlenecks can influence allele frequency patterns, large deviations from this gradient are unlikely. Consistent with these expectations, MAVE-aided stratification revealed a clear gradient, with allele frequencies decreasing across mild, intermediate, and damaging categories. VEP-aided stratification showed a broadly similar trend, though GEMME deviated from this pattern in three of the five genes, with some intermediate or damaging variants exhibiting discordant allele frequencies.

We next sought to investigate the variant classes in a protein structural context. Similar to the patterns observed with allele frequencies, we expected the different variant classes to be associated with distinct structural properties. For example, mild variants should be enriched in structurally tolerated amino acid substitutions located at protein surfaces, whereas damaging variants should be overrepresented in structurally perturbing substitutions mapping to buried or interface residues. To test this, we analysed high-quality, experimentally determined protein structures of the five cancer-associated genes and compared how these structural trends differed between MAVE- and VEP-aided stratification approaches.

In **Fig. 3A-B**, we examine the structural locations of residues affected by variants, grouped as interior, surface, or interface. As expected, both MAVE and VEP classifications reveal a clear trend: mild variants are more frequently associated with surface regions, while intermediate and damaging variants show an increasing propensity to localise in the protein interior. Consistent with previous studies [33–35], the uniquely tumour suppressor genes *BRCA1*, *MSH2*, and *PTEN* show a stronger enrichment of damaging variants in interior regions (see **Fig. S1** for gene-specific distributions), reflecting the importance of maintaining core structural integrity in these proteins [36]. In contrast, *HRAS* and *TP53*, which have oncogenic roles, exhibit a higher proportion of damaging variants at interfaces, highlighting the role of these regions in mediating gain-of-function effects such as abnormal signalling or transcription [37,38]. Although the overall patterns are similar, MAVE-aided stratification assigns a higher proportion of interface variants in *HRAS* to the damaging class compared to VEPs. Interestingly, this trend is also observed in the mild class, suggesting that not all interface variants are inherently damaging, but that MAVE scores are better able to distinguish between mild and damaging mutations.

**Fig. 3.**
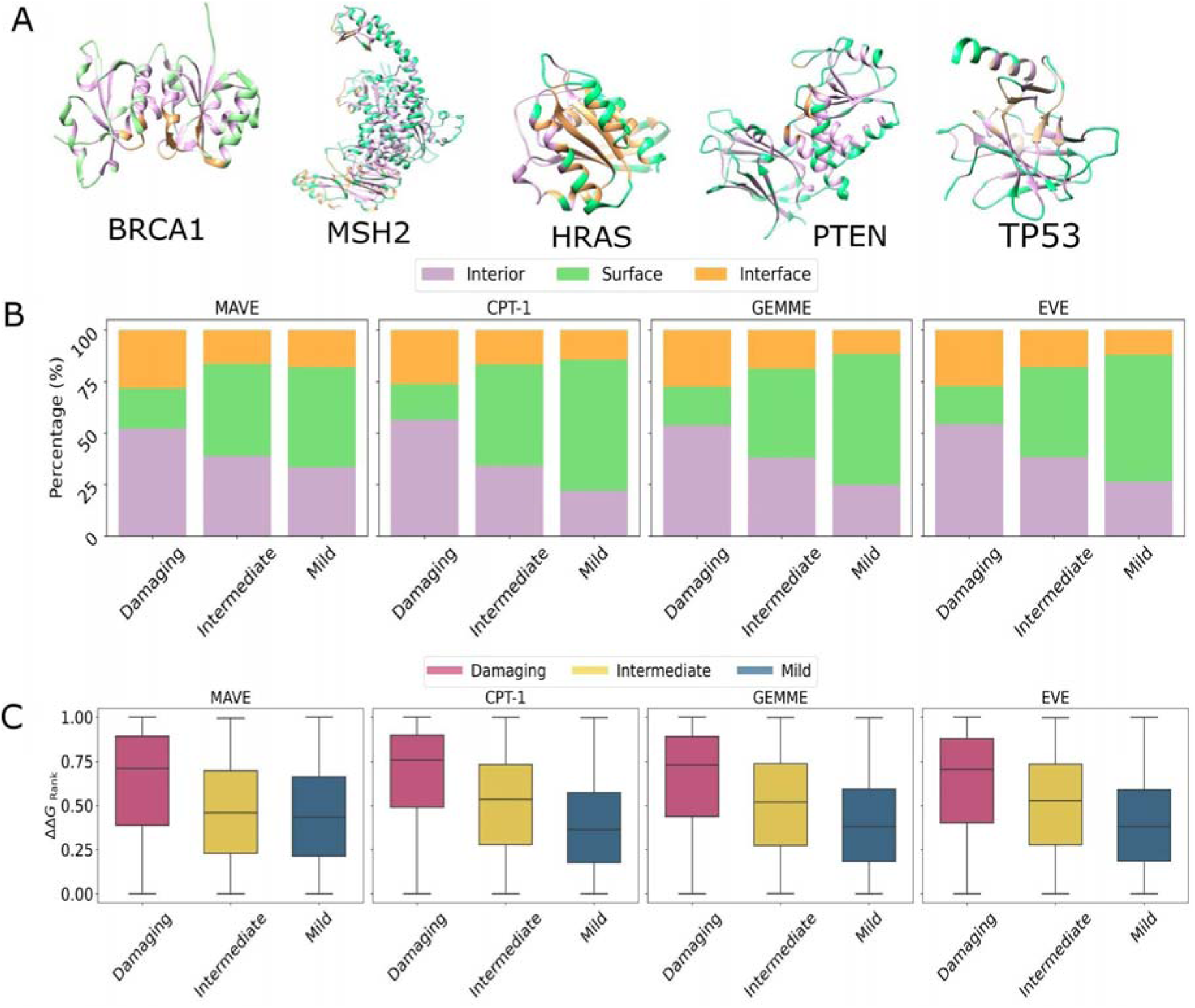
Structural properties of variants with MAVE- and VEP-aided stratification. **A:** Protein structures of the five genes coloured by residue location, *i.e.*, interior, surface, and interface. Structures represent the following PDB codes: pdb_00004y2g (BRCA1), pdb_00001wq1 (HRAS), pdb_00002o8b (MSH2), pdb_00001d5r (PTEN), and pdb_00005mct (*TP53*). **B:** Residue location composition of damaging, intermediate, and mild groups across the five proteins, grouped by MAVE, CPT-1, GEMME and EVE. **C:** ΔΔG_rank_ distribution of missense variants across damaging, intermediate, and mild categories across the five proteins. Boxes represent data within the 25th and 75th percentiles, the middle line is the median and the whiskers extend to 1.5× the interquartile range.

Next, we considered the variant classes in terms of their energetic impact within the structures. We used ΔΔG_rank_, a rank-normalised measure of the Gibbs free energy of folding (ΔΔG), with 0 representing the least and 1 the most structurally damaging variant in the protein [39]. In **Fig. 3C**, we show the ΔΔG_rank_ distribution of the different variant classes across genes. Similar to the trends we observed with the residue location data, the structural stability associated with the mild, intermediate, and damaging variants is largely similar for MAVE and VEPs. A notable exception is *HRAS*, where damaging variants as classified by the MAVE data have much milder structural effects compared to VEPs (see **Fig. S2** for gene-specific distributions). This is consistent with the gain-of-function molecular mechanism underlying *HRAS* disease phenotypes.

In summary, aside from assay-specific differences in HRAS, both MAVE- and VEP-aided stratification recover the expected biological gradients, with intermediate variants showing intermediate allele frequencies and structural effects. Contrary to our initial hypothesis, MAVEs did not consistently provide clearer separation of intermediate-impact variants than population-free VEPs. Under our chosen calibration, MAVEs classify fewer variants as damaging, suggesting a more conservative assignment, though this conservativeness reflects the applied thresholds and assay contexts and may vary with calibration choice and gene-specific mechanisms.

### ACMG-calibrated MAVEs yield fewer pathogenic and indeterminate classifications than VEPs

The MAVE- and VEP-aided stratification approach primarily serves to explore the structural and functional properties of variants; it is not intended for clinical classification purposes. Thus, we next examined if the observed differences would hold for clinical classification after calibrating MAVEs and VEPs to ACMG/AMP evidence strength [3,40,41]. Although this method was originally developed using variants aggregated across genes [5], the ClinGen Sequence Variant Interpretation Working Group recommends that functional data calibration be conducted at the gene level [4]. This is reinforced by a recent study highlighting potential pitfalls in genome-wide calibration [42], underscoring the need for gene-specific approaches. Such approaches offer key advantages, including the ability to account for genetic inheritance patterns and recognising that VEPs may exhibit heterogeneous distributions across different genes [43]. Consequently, thresholds determined from genome-wide data may not align well with the unique characteristics of individual genes, potentially leading to misclassification when applying the ACMG/AMP evidence thresholds.

To achieve gene-level calibration, we applied a kernel density estimation method, similar to previously established frameworks for VEP calibration [44,45], using score distributions from known pathogenic and (putatively) benign variants as reference to estimate positive likelihood ratios for each score value (**Fig. S3**) [41]. These likelihood ratios were then mapped onto ACMG/AMP evidence strengths, yielding clinically interpretable classifications for direct comparison between MAVE- and VEP-based evidence.

Two key observations emerge from the gene-level ACMG/AMP classification results presented in **Fig. 4**. First, calibrated MAVEs tend to yield the smallest fraction of variants reaching at least supporting level of evidence for pathogenicity in all genes except *BRCA1*. While this is consistent with the fraction of damaging variants in our stratification approach (**Fig. S1**), for *BRCA1* in particular, the two approaches seem to disagree. This likely stems from methodological differences: our stratification is based on a single ROC threshold, whereas the ACMG/AMP calibration considers the probability density along the entire score distribution (**Fig. S3**) [41]. This illustrates how methodological choices in score interpretation can substantially influence the resulting classification, emphasising the need for careful calibration when applying functional scores in clinical contexts.

**Fig. 4.**
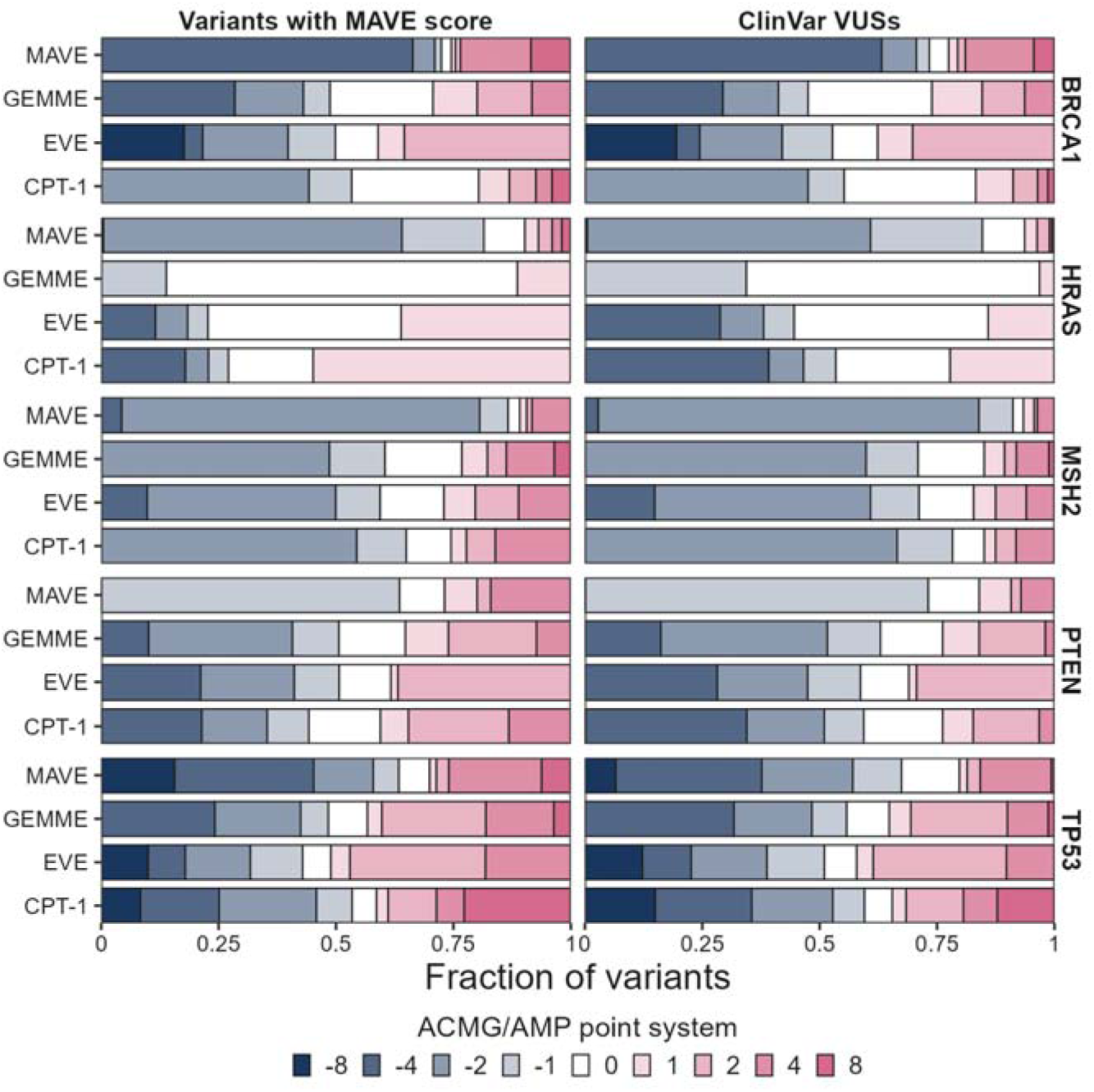
Missense variant classification using MAVE and VEP scores calibrated to ACMG/AMP evidence strength. Stacks show the fraction of variants reaching the given ACMG/AMP evidence level. Classifications are shown for all variants with MAVE scores and variants of uncertain significance (VUSs) in ClinVar. Colours correspond to the ACMG/AMP point system: benign very strong (-8), benign strong (-4), benign moderate (-2), benign supportive (-1), indeterminate (0) pathogenic supportive (1), pathogenic moderate (2), pathogenic strong (4), pathogenic very strong (8).

The second key observation is that calibrated MAVEs tend to yield the smallest proportion of variants classified as having indeterminate evidence. Although *TP53* was an exception, where CPT-1 produced a marginally lower indeterminate fraction, this trend held across the remaining genes. Of the 35,733 variants with MAVE coverage, only 1,845 (5.2%) were classified as indeterminate with respect to pathogenicity or benignity. In contrast, the corresponding figures were considerably higher for VEPs: 7,035 (19.7%) for GEMME, 4,557 (12.8%) for EVE, and 4,048 (11.3%) for CPT-1.

Consistent with this trend, and reflecting both lower pathogenic and indeterminate rates, calibrated MAVEs yielded the highest fraction of variants classified as benign, *i.e.*, variants reaching at least supporting-level evidence for benignity. Specifically, 27,293 (76.4%) MAVE-scored variants were classified as benign, compared to 18,379 (51.4%) for GEMME, 16,480 (46.1%) for EVE, and 19,593 (54.8%) for CPT-1. Notably, these trends held across ClinVar VUS (**Fig. 4**).

It is important to clarify that the *intermediate* and *indeterminate* classifications serve distinct purposes and are not directly comparable. Intermediate classifications represent the quartile of mild variants closest in effect to those classified as damaging. By contrast, in the ACMG/AMP framework, indeterminate variants are those that do not reach supporting evidence threshold for pathogenicity or benignity, *i.e.*, odds of pathogenicity of 2.08 or 0.481, respectively, under a prior probability of 0.1 for the gene. This distinction explains the apparent contradiction that MAVE-aided stratification tends to yield the largest fraction of intermediate variants, yet the smallest fraction of indeterminate variants when calibrated to ACMG/AMP evidence strength.

Taken together, our results from both the stratification and the ACMG/AMP calibration point to two distinctive characteristics of MAVE-based classification. First, it is more conservative: MAVE scores tend to assign fewer variants to the damaging or pathogenic category, even when predictive performance is strong. Second, it is more decisive: MAVE scores more clearly separate variants into benign or pathogenic, leading to fewer indeterminate calls. These properties are especially valuable in clinical settings, where overly permissive or uncertain classifications can complicate variant interpretation and prioritisation.

### Cross-population reclassification and prioritisation of variants with MAVEs and VEPs

The ability of MAVEs and population-free VEPs to classify variants independently of allele frequency makes them particularly useful for variant interpretation in diverse and historically underrepresented populations. These methods have demonstrated stable performance across ancestry groups and can help overcome limitations of clinical-trained and population-tuned VEPs, which rely heavily on European-skewed data [9]. Indeed, recent studies have shown that MAVEs can reclassify VUSs at higher rates in non-European populations [8], at a time when clinical sequencing continues to deliver substantially lower diagnostic yields for individuals of non-European ancestry [46].

Here, we extend this line of work by assessing missense variants in *BRCA1*, *HRAS*, *MSH2*, *PTEN*, and *TP53* observed in the SG10K_Health and Mexico City Prospective Study (MCPS) reference datasets. While strongly deleterious alleles in penetrant monogenic disorders are expected to be rare but broadly distributed across ancestry groups due to purifying selection [47], these five hereditary cancer genes provide ideal models for cross-population analysis. The presence of a variant in a population indicates that it is compatible with life, yet in the context of cancer predisposition genes, such variants may still confer elevated disease risk. Importantly, our use of population data is not aimed at identifying ancestry-specific risk alleles, but rather at expanding the discovery space for clinically actionable variants. Prioritisation with MAVEs can complement VEPs by detecting functional effects not fully captured by current models, with both approaches contributing to more equitable variant curation.

We combined evidence from ACMG/AMP-calibrated MAVEs and VEPs to reclassify variants from ClinVar that had been labelled as VUS, lacked clinical significance, or were unclassified entirely but observed in the SG10K_Health and MCPS datasets (N = 483). For each variant, we paired its MAVE-based ACMG/AMP classification with that of CPT-1, the best-performing VEP overall across the five genes. These were converted into evidence points, summed, and interpreted according to the Bayesian point system for combining evidence [48] following CanVIG-UK consensus recommendations [49]. We recognise that functional and computational evidence may not be fully independent, and it has been previously recommended to limit the combined weight of certain partially overlapping codes (e.g. PP3 with PM1) [5]. While no formal capping rule currently exists for PS3 with PP3, this remains an area of active discussion and should be considered in future guideline refinements.

Across the two cohorts, this approach reclassified 442 of 483 variants (91.5%) as benign or pathogenic, with the vast majority, 420 variants (87%), falling into the Benign and Likely Benign classes, and only 22 (4.6%) classified as Pathogenic or Likely Pathogenic; the remaining 41 (8.5%) retained the VUS classification (**Fig. 5A**). The 41 unresolved variants are visualised in a heatmap (**Fig. 5B**), illustrating that classification discrepancies typically stem from discordant MAVE and VEP evidence, with a tendency for VEPs to agree with each other. Of the 22 variants reclassified as Pathogenic or Likely Pathogenic, only one, A1789T in *BRCA1*, was observed in SG10K_Health, while the remaining 21 were found in the MCPS cohort. These include two variants in *BRCA1*, four in *MSH2*, and sixteen in *TP53*. All reclassified variants are listed in **Table S2**.

**Fig. 5.**
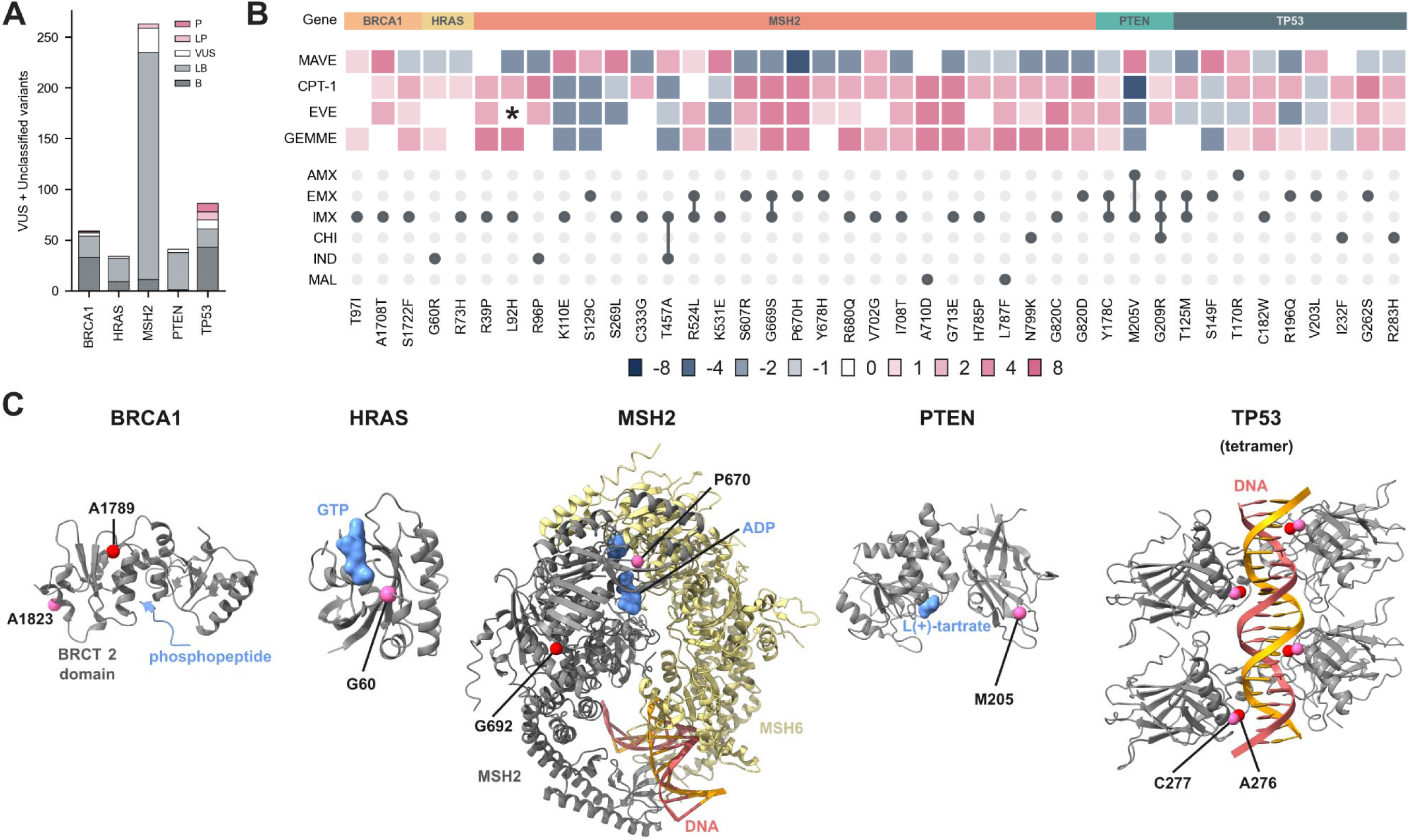
Missense variant reclassification and the structural context of pathogenic and discordant variants. **A**: Stacked bar chart showing the absolute counts of reclassified variants across the five genes from the MCPS and SG10K_Health cohorts. These variants were originally labelled in ClinVar as variants of uncertain significance (VUS) or carried no classification. Reclassification was performed using ACMG/AMP-calibrated MAVE and CPT-1 scores, with the positive likelihood ratios converted into the point-based evidence strengths, and summed. Final classifications (P, LP, VUS, LB, or B) were assigned according to ClinGen SVI recommendations. **B**: Heatmap displaying the subset of VUS or unclassified variants that remain VUS after reclassification based on combined MAVE and CPT-1 evidence. A star (*) indicates a variant with a missing EVE score. Colour shading reflects ACMG/AMP point-based strengths: benign very strong (-8), benign strong (-4), benign moderate (-2), benign supportive (-1), indeterminate (0), pathogenic supportive (+1), pathogenic moderate (+2), pathogenic strong (+4), and pathogenic very strong (+8). The accompanying UpSet plot indicates the ancestry groups in which each variant has been observed (AMX – African, EMX – European, IMX – Indigenous Mexican, CHI – Chinese, IND – Indian, MAL – Malay). **C**: Selected variants of uncertain significance classified as pathogenic or likely pathogenic by combining evidence from MAVEs and CPT-1 are shown in red, and those with discordant evidence are shown in pink (see **Table 2**). Structures represent the following PDB codes: pdb_00004y2g (*BRCA1*), pdb_00001wq1 (*HRAS*), pdb_00002o8b (*MSH2*), pdb_00001d5r (*PTEN*), and pdb_00005mct (*TP53*).

**Table 2.** Discordant variants across SG10K_Health and MCPS cohorts, where MAVE- and CPT-1-based ACMG/AMP calibrations yield conflicting evidence. MAVE and CPT-1 columns represent the ACMG/AMP evidence points: benign very strong (-8), benign strong (-4), benign moderate (-2), benign supportive (-1), indeterminate (0) pathogenic supportive (1), pathogenic moderate (2), pathogenic strong (4), pathogenic very strong (8). Variants in bold are discussed in the main text.

| Cohort | Gene | Variant | MAVE | CPT-1 | ClinVar classification |
| --- | --- | --- | --- | --- | --- |
| SG10K_Health | *HRAS* | G60R | -1 | 1 | Uncertain_significance |
| SG10K_Health | *MSH2* | R96P | -2 | 4 | Uncertain_significance |
| SG10K_Health | *MSH2* | T457A | 2 | -2 |  |
| SG10K_Health | *MSH2* | L787F | -1 | 2 | Uncertain_significance |
| SG10K_Health | *MSH2* | N799K | -2 | 2 | Uncertain_significance |
| SG10K_Health | *MSH2* | I904V | 1 | -2 |  |
| SG10K_Health | *PTEN* | G209R | -1 | 1 |  |
| SG10K_Health | *TP53* | R175C | -4 | 1 | Uncertain_significance |
| SG10K_Health | *TP53* | S269I | -4 | 4 | Likely_pathogenic |
| SG10K_Health | *TP53* | R283H | -1 | 2 | Uncertain_significance |
| SG10K_Health | *TP53* | P295L | 1 | -8 |  |
| MCPS | *BRCA1* | R71K | 8 | -2 | Pathogenic |
| MCPS | *BRCA1* | S1722F | -1 | 2 |  |
| MCPS | *BRCA1* | D1778N | 1 | -1 | Pathogenic/Likely_pathogenic |
| MCPS | *BRCA1* | D1818G | 8 | -2 | Pathogenic/Likely_pathogenic |
| MCPS | *BRCA1* | **A1823T** | 8 | -2 | Pathogenic/Likely_pathogenic |
| MCPS | *HRAS* | T35A | -2 | 1 | Uncertain_significance |
| MCPS | *HRAS* | I46T | -2 | 1 |  |
| MCPS | *HRAS* | A59V | -2 | 1 |  |
| MCPS | *HRAS* | **G60D** | -2 | 1 | Pathogenic |
| MCPS | *HRAS* | R73H | -1 | 1 | Uncertain_significance |
| MCPS | *HRAS* | E143K | -2 | 1 | Uncertain_significance |
| MCPS | *MSH2* | L92H | -2 | 2 |  |
| MCPS | *MSH2* | K110E | 4 | -2 | Uncertain_significance |
| MCPS | *MSH2* | S129C | 2 | -2 |  |
| MCPS | *MSH2* | E132K | -2 | 1 |  |
| MCPS | *MSH2* | S269L | 4 | -1 |  |
| MCPS | *MSH2* | C333G | -2 | 2 | Uncertain_significance |
| MCPS | *MSH2* | K531E | 4 | -1 | Uncertain_significance |
| MCPS | *MSH2* | S607R | -2 | 4 | Uncertain_significance |
| MCPS | *MSH2* | G669S | -2 | 4 | Uncertain_significance |
| MCPS | *MSH2* | **P670H** | -4 | 4 |  |
| MCPS | *MSH2* | P670T | -4 | 2 | Uncertain_significance |
| MCPS | *MSH2* | Y678C | -2 | 1 | Uncertain_significance |
| MCPS | *MSH2* | Y678H | -2 | 2 |  |
| MCPS | *MSH2* | R680Q | -2 | 2 | Uncertain_significance |
| MCPS | *MSH2* | I708T | -2 | 2 |  |
| MCPS | *MSH2* | G713E | -2 | 4 |  |
| MCPS | *MSH2* | M726T | -2 | 1 | Uncertain_significance |
| MCPS | *MSH2* | H785P | -1 | 2 |  |
| MCPS | *MSH2* | G820C | -2 | 2 | Uncertain_significance |
| MCPS | *MSH2* | G820D | -2 | 4 | Uncertain_significance |
| MCPS | *PTEN* | H93R | -1 | 2 | Pathogenic |
| MCPS | *PTEN* | T167N | -1 | 2 | Likely_pathogenic |
| MCPS | *PTEN* | Y178C | -1 | 1 |  |
| MCPS | *PTEN* | **M205V** | 4 | -4 | Uncertain_significance |
| MCPS | *PTEN* | Q396P | 2 | -4 |  |
| MCPS | *TP53* | T125M | -2 | 2 |  |
| MCPS | *TP53* | S149F | 4 | -1 | Uncertain_significance |
| MCPS | *TP53* | R175P | -1 | 8 |  |
| MCPS | *TP53* | C182W | -1 | 2 | Uncertain_significance |
| MCPS | *TP53* | R196Q | -2 | 2 | Uncertain_significance |
| MCPS | *TP53* | V203L | 2 | -1 | Uncertain_significance |
| MCPS | *TP53* | G262S | -1 | 4 | Uncertain_significance |
| MCPS | *TP53* | **C277R** | -2 | 8 |  |
| MCPS | *TP53* | R337C | 4 | -2 | Pathogenic/Likely_pathogenic |
| MCPS | *TP53* | A347D | -4 | 4 | Pathogenic |

We highlight one pathogenic (reclassified) variant from each of the three genes, with residue positions shown in structural context in **Fig. 5C**. The *BRCA1* variant A1789T is observed in one individual of South Asian ancestry in gnomAD and one individual of Indian ancestry in SG10K_Health. Located within the *BRCT* 2 domain, this variant has a calibrated MAVE score indicating strong evidence for pathogenicity, with CPT-1 contributing moderate pathogenic evidence (4+2=6 points; Likely Pathogenic). Functional characterisation across multiple assays supports a deleterious effect on *BRCA1*-mediated DNA repair. The variant significantly impairs non-homologous end-joining activity in HeLa cells and alters double-strand break repair [50], while in yeast it disrupts growth suppression, consistent with loss of tumour suppressor function [51,52]. Expression of A1789T in human cells perturbs the transcription of hundreds of genes involved in DNA damage response and cell cycle progression [53], and the variant fails to rescue cisplatin sensitivity in *BRCA1*-deficient cells [54], reinforcing its loss of function across multiple DNA repair contexts.

The *MSH2* variant G692E is absent from gnomAD and SG10K_Health, but present in one individual of European ancestry in MCPS. It lies within the ATPase domain of MSH2, near the base of a conserved helix that contributes to nucleotide binding and conformational coupling [55]. The calibrated MAVE score of the variant indicates strong evidence for pathogenicity, with CPT-1 contributing strong pathogenicity evidence (4+4=8 points; Likely Pathogenic). Functional testing from a yeast-based complementation assay indicates that G692E substantially elevates the mutation rate, consistent with loss of mismatch repair activity [56]. Interestingly, other substitutions at the same residue (G692R, G692W, and G692V) are classified as (likely) pathogenic in ClinVar, in association with cancer predisposition syndromes, further supporting the clinical significance of this variant.

The *TP53* variant A276D has a single allele in gnomAD but sixty-two in MCPS, mainly in individuals of indigenous Mexican and European ancestry. Functional characterisation across multiple assay systems consistently supports its loss of function. The calibrated MAVE score of the variant indicates strong evidence for pathogenicity, with CPT-1 contributing very strong pathogenicity evidence (4+8=12 points; Pathogenic). In mammalian cells, A276D retains less than 10% of wild-type transactivation activity on an MDM2 reporter and fails to suppress the growth of *TP53*-null H1299 cells, while also lacking growth-suppressive activity in a yeast colony assay [57]. It further shows near-complete loss of transactivation capacity across a panel of TP53-responsive promoters in a yeast-based luciferase reporter assay [58]. Additionally, A276D fails to repress Nrf2 promoter activity in H1299 cells, in contrast to wild-type *TP53*, which strongly suppresses Nrf2 expression at both protein and reporter levels [59], reinforcing its loss of transcriptional function.

### Variants with discordant evidence highlight method-specific limitations

While MAVE and VEP evidence were generally concordant, allowing us to reclassify over 90% of VUS and unclassified variants, we examined those cases where the two sources provided opposing evidence and effectively cancelled each other out: one supporting pathogenicity, the other benign classification. Variants with indeterminate scores from either source were excluded from this analysis. We identified sixty-five such discordant variants across the five genes in the SG10K_Health and MCPS cohorts, listed in **Table 2**. Here, we discuss representative variants from each gene, with residue positions visualised in **Fig. 5C**.

A missense variant in BRCA1, also recently discussed [60], was A1823T, located within the BRCT2 domain. This variant has only been observed once in an individual of indigenous Mexican ancestry. The MAVE assay classified the variant as pathogenic very strong (8), while CPT-1 assigned moderate benign evidence (-2). Recently, Adamovich et al. noted that A1823T falls at a splice junction and is associated with reduced mRNA expression [61], consistent with a high SpliceAI score of 0.65 predicting aberrant splicing [62]. CPT-1 is unlikely to detect splice-disruptive effects unless the affected residue is also evolutionarily constrained. In contrast, the MAVE assay employs an endogenous construct in human cells and is sensitive to missense changes that disrupt splicing. Based on the available evidence, A1823T likely represents a risk allele.

The *HRAS* variant G60R also exhibited discordant evidence. The MAVE assay classified G60R as moderately benign (-2), whereas CPT-1 contributed supporting pathogenic evidence (+1). It has been observed once among 3,552 Japanese individuals [63] and once in gnomAD. A patient carrying G60R was diagnosed with Costello syndrome, although the variant alone was considered unlikely to fully explain their phenotype [64]. Functional studies indicate that the G60R mutant HRAS binds more strongly to RAF1 than the wild type, suggesting altered downstream signalling [65]. Other substitutions at this residue (G60V, G60D, and G60R) are classified as (likely) pathogenic in ClinVar, supporting the functional importance of this position. While the MAVE assay has outstanding performance in HRAS, particularly for gain-of-function variants and those affecting GTP binding, it was not designed to capture downstream effects on MAPK signalling that depend on human-specific protein interactions, such as enhanced RAF1 binding. Likewise, if the molecular mechanism involves gain of function, VEPs, which generally perform better on loss-of-function variants, are expected to underperform [30]. Based on the available evidence, it cannot be excluded that G60R represents a risk allele.

The *MSH2* variant P670H presents a third case of discordant evidence. It is absent from gnomAD and SG10K_Health but was identified in four individuals of European ancestry in the MCPS cohort. Located within the ATPase domain of MSH2, this residue is in direct contact with ATP (**Fig. 5C**) [66]. The MAVE assay classified P670H as strongly benign (-4), whereas CPT-1 provided strong pathogenic evidence (+4), effectively cancelling each other. The variant has previously been reported in individuals with breast and colorectal cancer [67,68]. Given the non-conservative nature of the proline-to-histidine substitution, the residue’s proximity to the ATP-binding site, and the fact that two other variants at this position, S607R and S607T, are also discordant (**Table 2**), it is plausible that VEPs like CPT-1 may be overly sensitive to positional conservation. In contrast, the MAVE assay, which is highly sensitive to loss of function, found no functional impact, suggesting that the variant does not impair ATP binding or hydrolysis in the cellular context. Whether P670H contributes to cancer risk remains unresolved.

The *PTEN* variant M205V received discordant classification, with strong pathogenic evidence from the MAVE assay (+4) counterbalanced by strong benign evidence from CPT-1 (-4). M205V is currently classified as a VUS in ClinVar and has been observed nineteen times in gnomAD. It was also found in three individuals from the MCPS cohort: one of indigenous Mexican ancestry and two of African ancestry. A VAMP-seq assay classified the variant as ‘possibly low’ in protein abundance, suggestive of impaired stability [69]. M205V has been reported in Cowden syndrome cohorts [70,71], but patient-level data provide little support for a strong pathogenic effect as the variant was not observed in any female breast cancer cases and appeared once among control cases [72]. From a structural perspective, small-angle X-ray scattering data indicate that M205V does not affect the PTEN dimer interface [73]. Taken together, these findings suggest that while M205V may modestly reduce PTEN abundance and phosphatase activity, its clinical relevance remains uncertain, and a risk-allele interpretation cannot be excluded.

The *TP53* variant C277R was discordant, with the MAVE assay classifying it as moderately benign (-2), while CPT-1 contributed very strong pathogenic evidence (+8). It lies adjacent to A276D, a variant of uncertain significance that we reclassified as pathogenic (**Fig. 5C**), suggesting it may share a molecular mechanism. Functional studies consistently show that C277R disrupts DNA binding, reducing both affinity and specificity [74,75]. It is defective in CDKN1A induction [76] and exhibits impaired transactivation capacity in multiple assays [58,77]. Expression under the GAL1 promoter caused growth inhibition comparable to a double mutant [78]. C277R was identified in a patient with early-onset breast cancer and a family history of breast and prostate cancers [79], and has been observed in glioma and carcinoma samples [80]. A combined ACMG/AMP and somatic-to-germline ratio analysis yielded a high posterior probability of pathogenicity (0.946) [81]. Collectively, these data support the CPT-1 classification that C277R is likely pathogenic.

### Conclusions

This comparative analysis of MAVEs and VEPs across five cancer-associated genes reveals distinct yet complementary strengths in variant classification properties. Our systematic evaluation demonstrates that MAVEs exhibit more conservative classification behaviour, consistently assigning fewer variants to damaging or pathogenic categories while maintaining strong predictive performance. ACMG/AMP-calibrated MAVEs are also more decisive, yielding substantially fewer indeterminate classifications compared to population-free VEPs. This shift towards fewer pathogenic and indeterminate calls occurs even though both experimental and computational approaches still produce biologically coherent stratifications. Intermediate-effect variants largely follow the expected allele frequency gradient and structural patterns. Although MAVEs provide clear, context-dependent functional readouts, VEPs draw upon evolutionary conservation to capture a broader range of molecular consequences, such as effects on cryptic interface residues that fall outside the scope of the assay, thereby complementing areas where MAVEs may under-sample. These qualities minimise the burden of false-positive or ambiguous findings in clinical workflows, allowing resources to be focused on variants with the highest confidence for follow-up assays and patient management. The clinical utility of combining MAVE and VEP evidence is exemplified by the reclassification of over 90% of variants of uncertain significance in ancestrally diverse cohorts, transforming previously ambiguous results into definitive benign or pathogenic classifications and thereby enabling robust and equitable genetic interpretation. As the field advances towards precision medicine, exploiting such complementary approaches will be essential to translate genomic insights into meaningful patient care for all.

## Methods

### Sources of missense variation

We collected missense variants from the following databases: ClinVar (2025-05-06 release) [24], gnomAD v4.1 [82], SG10K_Health [21,22], Mexico City Prospective Study (MCPS) [23]. For both P/LP and B/LB classes, we included only variants with a ClinVar review status of at least one star. Variants in SG10K_Health and MCPS were processed as previously described [9]. For the other sources, we processed variant calling files with GRCh38 coordinates using bcftools 1.20 [83] and mapped them to UniProt reference sequences via an in-house pipeline based on Ensembl VEP 112 [84].

### AUROC and optimal classification threshold

The area under the receiver operating characteristic (AUROC) curve was calculated in R using the roc() function from the pROC package v1.18.5. The optimal threshold, defined as the distance from the (0,1) point of the ROC curve, was estimated with the pROC::coords() function by selecting the ‘closest.topleft’ option for the best.method argument.

### MAVE and VEP scores

As our interest was to compare VEPs to the clinically most predictive MAVE study in all five cancer genes, we selected the fitness map with the highest AUROC in a binary classification task performed on missense pathogenic and benign variants from ClinVar or missense putatively benign variants from gnomAD in the case of *HRAS* and *PTEN*. This process resulted in the following fitness maps (gene name -original fitness score name): *BRCA1* -function.score.mean [14]; *HRAS* -DMS_attenuated [17]; *MSH2* -LOF score [18]; *TP53* -A549_p53NULL_Etoposide_Z-score [20]; *PTEN* -Cum_score [19]. GEMME and CPT-1 scores were complete for the variants covered by the fitness maps (N = 35,733). EVE had 8.8% missing values, with the highest rate observed in *TP53* at 16.3%.

### Allele frequency calculation

Allele frequencies depicted in **Fig. 2C** were calculated using exome data from gnomAD v4.1. Variants from the Ashkenazi Jewish (ASJ) and Finnish (FIN) ancestry groups were excluded, as these relatively isolated populations can harbour pathogenic variants at higher frequencies than more admixed populations. The analysis therefore included only variants from the African/African American (AFR), Latino/Admixed American (AMR), East Asian (EAS), Middle Eastern (MID), non-Finnish European (NFE), and South Asian (SAS) ancestry groups. To allow representation on a logarithmic scale, we computed the geometric mean of cumulative allele frequencies across these six groups for the mild, intermediate, and damaging classes.

### Calibration to ACMG/AMP evidence strength

MAVE and VEP scores were calibrated under ACMG/AMP guidelines using the acmgscaler R package [41]. For all genes, we adopted the default prior probability of pathogenicity of 0.1, consistent with a previous global estimate [40]. This prior also underpins the CanVIG-UK consensus recommendations [49], which we followed when combining MAVE and VEP evidence. While we recognise that the true prior may vary between genes and even across disease phenotypes, estimating these priors remains a non-trivial task. Importantly, our focus was to compare the relative classification behaviour of MAVEs and VEPs, which is not dependent on the choice of the prior.

### Structural data

For structural analysis, we selected one representative structure from the Protein Data Bank (PDB) [85] for each of the five proteins, either based on the chain with the highest coverage of the UniProt sequence (*BRCA1*, *MSH2*, *PTEN*, and *TP53*) or its relevance to the condition of the fitness map (HRAS). The following structures (PDB identifiers) were selected: pdb_00004y2g (*BRCA1*), pdb_00001wq1 (*HRAS*), pdb_00002o8b (*MSH2*), pdb_00001d5r (*PTEN*), and pdb_00005mct (*TP53*). Solvent accessible surface area was calculated at residue level with AREAIMOL from the CCP4 suite [86]. These were normalised by the maximum surface area measured in Gly-X-Gly tripeptides [87] to assign residue locations [88]. The change in the Gibbs free energy (ΔΔG) for each variant was estimated with FoldX 5.0 [89], by first calling the RepairPDB command on the structure followed by the BuildModel command.

## Data Availability

All data produced in the present study are available upon reasonable request to the authors.

## Acknowledgements

The project was a part of the LKCMed-University of Edinburgh Collaboration (Grant no: WBS: 03INP001356A630). This study made use of data generated as part of the Singapore National Precision Medicine program funded by the Industry Alignment Fund (Pre-Positioning) (IAF-PP: H17/01/a0/007). This study made use of data/samples collected in the following cohorts in Singapore: (1) The Health for Life in Singapore (HELIOS) study at the Lee Kong Chian School of Medicine, Nanyang Technological University, Singapore (supported by grants from a Strategic Initiative at Lee Kong Chian School of Medicine, the Singapore Ministry of Health under its Singapore Translational Research Investigator Award (NMRC/STaR/0028/2017) and the IAF-PP: H18/01/a0/016); (2) The Growing up in Singapore Towards Healthy Outcomes (GUSTO) study, which is jointly hosted by the National University Hospital (NUH), KK Women’s and Children’s Hospital (KKH), the National University of Singapore (NUS) and the Singapore Institute for Clinical Sciences (SICS), Agency for Science Technology and Research (A*STAR) (supported by the Singapore National Research Foundation under its Translational and Clinical Research (TCR) Flagship Programme and administered by the Singapore Ministry of Health’s National Medical Research Council (NMRC), Singapore -NMRC/TCR/004-NUS/2008); (3) The Singapore Epidemiology of Eye Diseases (SEED) cohort at Singapore Eye Research Institute (SERI) (supported by NMRC/CIRG/1417/2015; NMRC/CIRG/1488/2018; NMRC/OFLCG/004/2018); (4) The Multi-Ethnic Cohort (MEC) cohort (supported by NMRC grant 0838/2004; BMRC grant 03/1/27/18/216; 05/1/21/19/425; 11/1/21/19/678, Ministry of Health, Singapore, National University of Singapore and National University Health System, Singapore); (5) The SingHealth Duke-NUS Institute of Precision Medicine (PRISM) cohort (supported by NMRC/CG/M006/2017_NHCS; NMRC/STaR/0011/2012, NMRC/STaR/ 0026/2015, Lee Foundation and Tanoto Foundation); (6) The TTSH Personalised Medicine Normal Controls (TTSH) cohort funded (supported by NMRC/CG12AUG17 and CGAug16M012).

The views expressed are those of the author(s) and are not necessarily those of the National Precision Medicine investigators, or institutional partners. We thank all investigators, staff members and study participants who made the National Precision Medicine Project possible.

This project was supported by funding to JAM from the European Research Council (ERC) under the European Union’s Horizon 2020 research and innovation programme (grant agreement No. 101001169), a Lister Institute Research Prize Fellowship and by the Medical Research Council (MRC) Human Genetics Unit core grant (MC_UU_00035/9). This project was supported by the PRECISE grants (NRPRECI221, NRPREWT231) and National Medical Research Council Singapore Clinician-Scientist Award (NMRC/CSA-INV/0017/2017, MOH000654) grant conducted at the National Cancer Centre Singapore to J.N.

## Supplemental figures

**Fig. S1.**
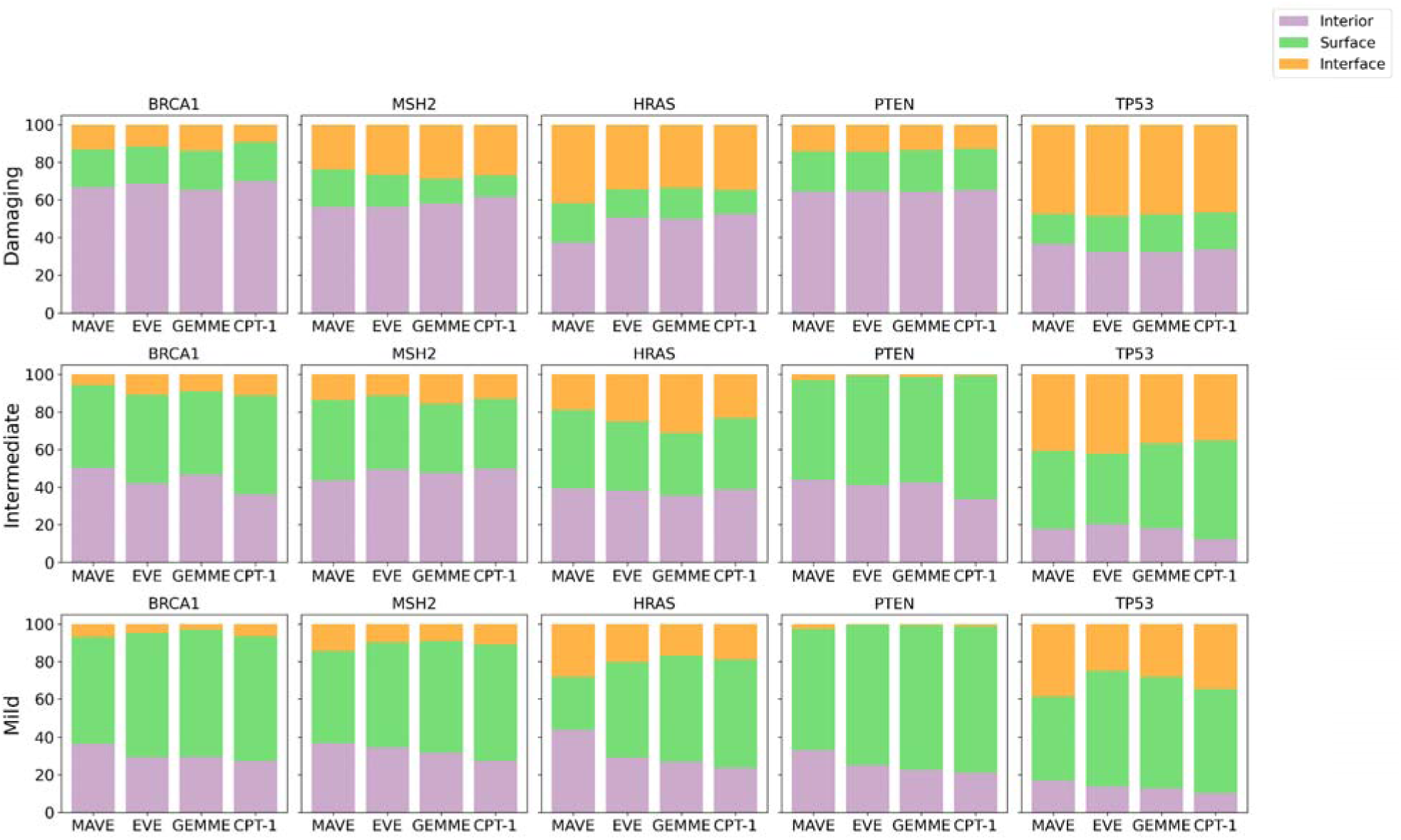
Residue location composition of damaging, intermediate, and mild groups separately for the five genes.

**Fig. S2.**
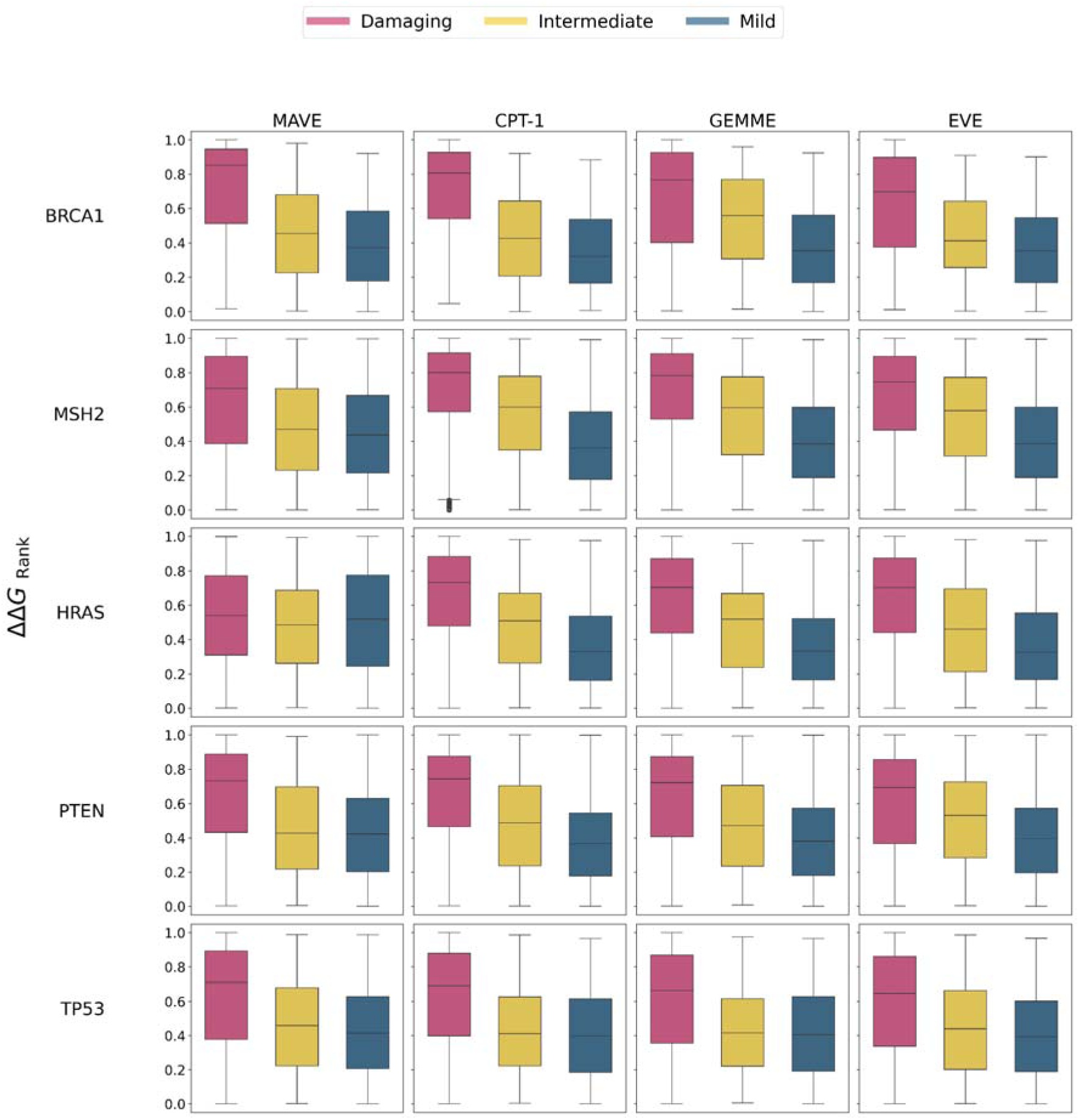
ΔΔGrank distribution of the missense variants for damaging, intermediate, and mild groups separately for the five genes. Boxes represent data within the 25th and 75th percentiles, the middle line is the median and the whiskers extend to 1.5× the interquartile range.

**Fig. S3.**
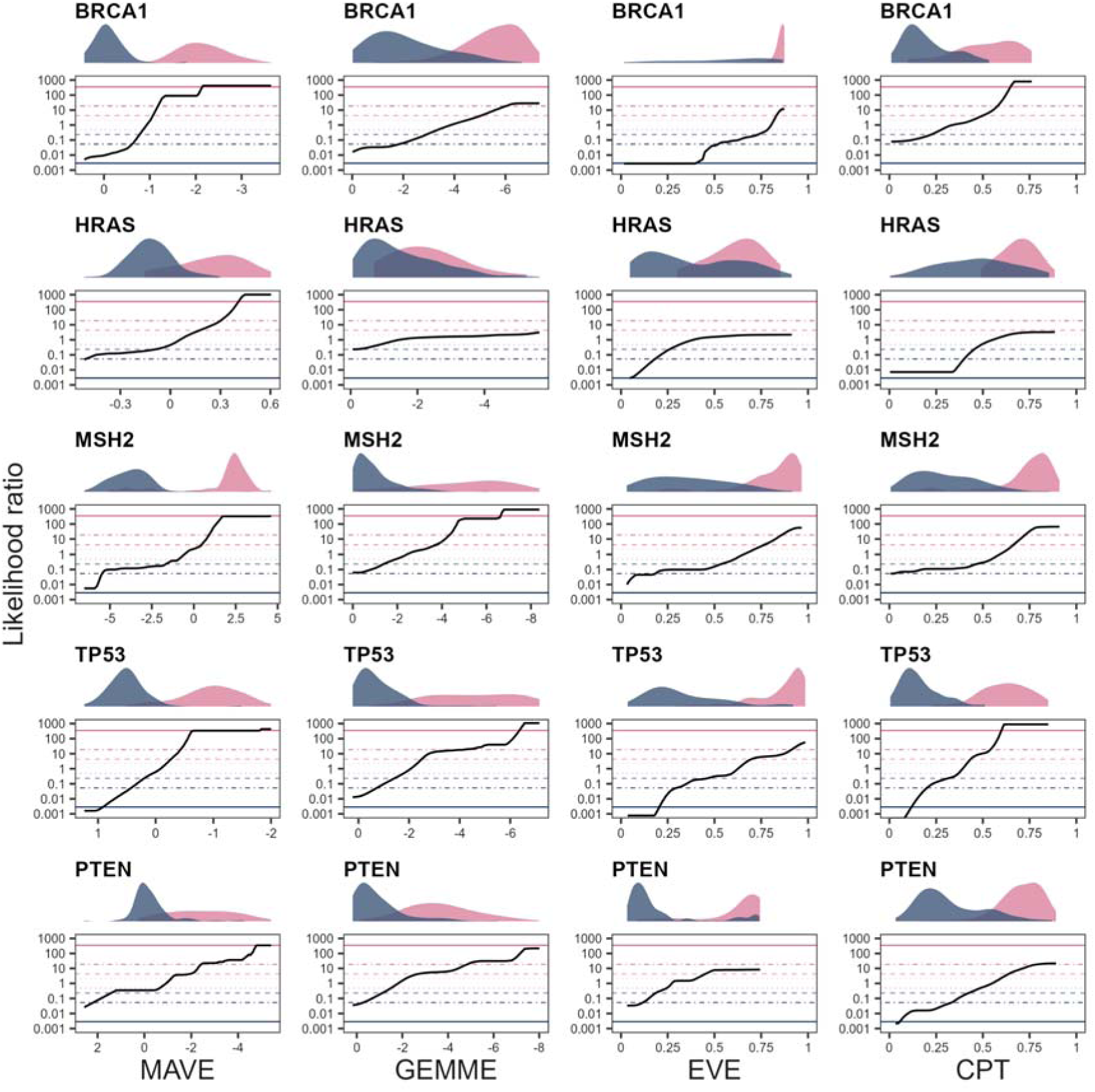
ACMG/AMP calibration of MAVE and VEP scores. Density plots show the distribution of pathogenic (pink) and benign (blue) scores. Horizontal bars in the likelihood ratio charts correspond to evidence strengths under a prior probability of pathogenicity of 0.1 (from top to bottom: pathogenic very strong, pathogenic strong, pathogenic moderate, pathogenic supportive, benign supportive, benign moderate, benign strong, benign very strong).

## References

1. Chen E, Facio FM, Aradhya KW, Rojahn S, Hatchell KE, Aguilar S, et al. Rates and Classification of Variants of Uncertain Significance in Hereditary Disease Genetic Testing. JAMA Netw Open. 2023;6:e2339571. 10.1001/jamanetworkopen.2023.39571

2. Kobayashi Y, Chen E, Facio FM, Metz H, Poll SR, Swartzlander D, et al. Clinical Variant Reclassification in Hereditary Disease Genetic Testing. JAMA Netw Open. 2024;7:e2444526. 10.1001/jamanetworkopen.2024.44526

3. Richards S, Aziz N, Bale S, Bick D, Das S, Gastier-Foster J, et al. Standards and guidelines for the interpretation of sequence variants: a joint consensus recommendation of the American College of Medical Genetics and Genomics and the Association for Molecular Pathology. Genetics in Medicine. Elsevier; 2015;17:405–24. 10.1038/gim.2015.30

4. Brnich SE, Abou Tayoun AN, Couch FJ, Cutting GR, Greenblatt MS, Heinen CD, et al. Recommendations for application of the functional evidence PS3/BS3 criterion using the ACMG/AMP sequence variant interpretation framework. Genome Med. 2019;12:3. 10.1186/s13073-019-0690-2

5. Pejaver V, Byrne AB, Feng B-J, Pagel KA, Mooney SD, Karchin R, et al. Calibration of computational tools for missense variant pathogenicity classification and ClinGen recommendations for PP3/BP4 criteria. The American Journal of Human Genetics. Elsevier; 2022;109:2163–77. 10.1016/j.ajhg.2022.10.013

6. Grimm DG, Azencott C-A, Aicheler F, Gieraths U, MacArthur DG, Samocha KE, et al. The Evaluation of Tools Used to Predict the Impact of Missense Variants Is Hindered by Two Types of Circularity. Human Mutation. 2015;36:513–23. 10.1002/humu.22768

7. Livesey BJ, Marsh JA. Variant effect predictor correlation with functional assays is reflective of clinical classification performance. Genome Biol. 2025;26:104. 10.1186/s13059-025-03575-w

8. Dawood M, Fayer S, Pendyala S, Post M, Kalra D, Patterson K, et al. Using multiplexed functional data to reduce variant classification inequities in underrepresented populations. Genome Med. 2024;16:143. 10.1186/s13073-024-01392-7

9. Pathak AK, Bora N, Badonyi M, Livesey BJ, Consortium S, Ngeow J, et al. Pervasive ancestry bias in variant effect predictors [Internet]. bioRxiv; 2024 [cited 2024 Aug 20]. p. 2024.05.20.594987. 10.1101/2024.05.20.594987

10. Tallman S, Moutsianas L, Nguyen T, Cho Y, Mackintosh M, Kasperaviciute D, et al. Missing genetic diversity impacts variant prioritisation for rare disorders [Internet]. medRxiv; 2024 [cited 2024 Aug 20]. p. 2024.08.12.24311664. 10.1101/2024.08.12.24311664

11. Fowler DM, Fields S. Deep mutational scanning: a new style of protein science. Nat Methods. Nature Publishing Group; 2014;11:801–7. 10.1038/nmeth.3027

12. Tabet D, Parikh V, Mali P, Roth FP, Claussnitzer M. Scalable Functional Assays for the Interpretation of Human Genetic Variation. Annual Review of Genetics. Annual Reviews; 2022;56:441–65. 10.1146/annurev-genet-072920-032107

13. Weile J, Roth FP. Multiplexed assays of variant effects contribute to a growing genotype–phenotype atlas. Hum Genet. 2018;137:665–78. 10.1007/s00439-018-1916-x

14. Findlay GM. Linking genome variants to disease: scalable approaches to test the functional impact of human mutations. Human Molecular Genetics. 2021;30:R187–97. 10.1093/hmg/ddab219

15. McEwen AE, Tejura M, Fayer S, Starita LM, Fowler DM. Multiplexed assays of variant effect for clinical variant interpretation. Nat Rev Genet. 2025; 10.1038/s41576-025-00870-x

16. Findlay GM, Daza RM, Martin B, Zhang MD, Leith AP, Gasperini M, et al. Accurate classification of BRCA1 variants with saturation genome editing. Nature. 2018;562:217–22. 10.1038/s41586-018-0461-z

17. Bandaru P, Shah NH, Bhattacharyya M, Barton JP, Kondo Y, Cofsky JC, et al. Deconstruction of the Ras switching cycle through saturation mutagenesis. Valencia A, editor. eLife. eLife Sciences Publications, Ltd; 2017;6:e27810. 10.7554/eLife.27810

18. Jia X, Burugula BB, Chen V, Lemons RM, Jayakody S, Maksutova M, et al. Massively parallel functional testing of MSH2 missense variants conferring Lynch syndrome risk. The American Journal of Human Genetics. Elsevier; 2021;108:163–75. 10.1016/j.ajhg.2020.12.003

19. Mighell TL, Evans-Dutson S, O’Roak BJ. A Saturation Mutagenesis Approach to Understanding PTEN Lipid Phosphatase Activity and Genotype-Phenotype Relationships. Am J Hum Genet. 2018;102:943–55. 10.1016/j.ajhg.2018.03.018

20. Giacomelli AO, Yang X, Lintner RE, McFarland JM, Duby M, Kim J, et al. Mutational processes shape the landscape of TP53 mutations in human cancer. Nat Genet. Nature Publishing Group; 2018;50:1381–7. 10.1038/s41588-018-0204-y

21. Chan SH, Bylstra Y, Teo JX, Kuan JL, Bertin N, Gonzalez-Porta M, et al. Analysis of clinically relevant variants from ancestrally diverse Asian genomes. Nat Commun. Nature Publishing Group; 2022;13:6694. 10.1038/s41467-022-34116-9

22. Wong E, Bertin N, Hebrard M, Tirado-Magallanes R, Bellis C, Lim WK, et al. The Singapore National Precision Medicine Strategy. Nat Genet. Nature Publishing Group; 2023;55:178–86. 10.1038/s41588-022-01274-x

23. Ziyatdinov A, Torres J, Alegre-Díaz J, Backman J, Mbatchou J, Turner M, et al. Genotyping, sequencing and analysis of 140,000 adults from Mexico City. Nature. Nature Publishing Group; 2023;622:784–93. 10.1038/s41586-023-06595-3

24. Landrum MJ, Lee JM, Benson M, Brown GR, Chao C, Chitipiralla S, et al. ClinVar: Improving access to variant interpretations and supporting evidence. Nucleic Acids Research. Nucleic Acids Res; 2018;46:D1062–7. 10.1093/nar/gkx1153

25. Chen S, Francioli LC, Goodrich JK, Collins RL, Kanai M, Wang Q, et al. A genomic mutational constraint map using variation in 76,156 human genomes. Nature. Nature Publishing Group; 2024;625:92–100. 10.1038/s41586-023-06045-0

26. Laine E, Karami Y, Carbone A. GEMME: A Simple and Fast Global Epistatic Model Predicting Mutational Effects. Molecular Biology and Evolution. 2019;36:2604–19. 10.1093/molbev/msz179

27. Frazer J, Notin P, Dias M, Gomez A, Min JK, Brock K, et al. Disease variant prediction with deep generative models of evolutionary data. Nature. Nature Publishing Group; 2021;599:91–5. 10.1038/s41586-021-04043-8

28. Jagota M, Ye C, Albors C, Rastogi R, Koehl A, Ioannidis N, et al. Cross-protein transfer learning substantially improves disease variant prediction. Genome Biol. 2023;24:182. 10.1186/s13059-023-03024-6

29. Meier J, Rao R, Verkuil R, Liu J, Sercu T, Rives A. Language models enable zero-shot prediction of the effects of mutations on protein function. Advances in Neural Information Processing Systems [Internet]. Curran Associates, Inc.; 2021 [cited 2025 May 5]. p. 29287–303. https://proceedings.neurips.cc/paper/2021/hash/f51338d736f95dd42427296047067694-Abstract.html. Accessed 5 May 2025

30. Gerasimavicius L, Livesey BJ, Marsh JA. Loss-of-function, gain-of-function and dominant-negative mutations have profoundly different effects on protein structure. Nature Communications 2022 13:1. Nature Publishing Group; 2022;13:1–15. 10.1038/s41467-022-31686-6

31. Gerasimavicius L, Teichmann SA, Marsh JA. Leveraging protein structural information to improve variant effect prediction. Curr Opin Struct Biol. 2025;92:103023. 10.1016/j.sbi.2025.103023

32. Yehia L, Ngeow J, Eng C. PTEN-opathies: from biological insights to evidence-based precision medicine. J Clin Invest. American Society for Clinical Investigation; 2019;129:452–64. 10.1172/JCI121277

33. Elze L, van der Post RS, Vos JR, Mensenkamp AR, Pamidimarri Naga S, Hampstead JE, et al. Genomic instability in non–breast or ovarian malignancies of individuals with germline pathogenic variants in BRCA1/2. JNCI: Journal of the National Cancer Institute. 2024;djae160. 10.1093/jnci/djae160

34. Bronner CE, Baker SM, Morrison PT, Warren G, Smith LG, Lescoe MK, et al. Mutation in the DNA mismatch repair gene homologue hMLH1 is associated with hereditary non-polyposis colon cancer. Nature. 1994;368:258–61. 10.1038/368258a0

35. Serebriiskii IG, Pavlov V, Tricarico R, Andrianov G, Nicolas E, Parker MI, et al. Comprehensive characterization of PTEN mutational profile in a series of 34,129 colorectal cancers. Nat Commun. Nature Publishing Group; 2022;13:1618. 10.1038/s41467-022-29227-2

36. Stehr H, Jang S-HJ, Duarte JM, Wierling C, Lehrach H, Lappe M, et al. The structural impact of cancer-associated missense mutations in oncogenes and tumor suppressors. Mol Cancer. 2011;10:54. 10.1186/1476-4598-10-54

37. Gripp KW, Stabley DL, Nicholson L, Hoffman JD, Sol-Church K. Somatic mosaicism for an HRAS mutation causes Costello syndrome. Am J Med Genet A. 2006;140:2163–9. 10.1002/ajmg.a.31456

38. Dittmer D, Pati S, Zambetti G, Chu S, Teresky AK, Moore M, et al. Gain of function mutations in p53. Nat Genet. Nature Publishing Group; 1993;4:42–6. 10.1038/ng0593-42

39. Chillón-Pino D, Badonyi M, Semple CA, Marsh JA. Protein structural context of cancer mutations reveals molecular mechanisms and candidate driver genes. Cell Rep. 2024;43:114905. 10.1016/j.celrep.2024.114905

40. Tavtigian SV, Greenblatt MS, Harrison SM, Nussbaum RL, Prabhu SA, Boucher KM, et al. Modeling the ACMG/AMP variant classification guidelines as a Bayesian classification framework. Genetics in Medicine. 2018;20:1054–60. 10.1038/gim.2017.210

41. Badonyi M, Marsh JA. acmgscaler: An R package and Colab for standardised gene-level variant effect score calibration within the ACMG/AMP framework. Bioinformatics. 2025;btaf503. 10.1093/bioinformatics/btaf503

42. Tejura M, Fayer S, McEwen AE, Flynn J, Starita LM, Fowler DM. Calibration of variant effect predictors on genome-wide data masks heterogeneous performance across genes. The American Journal of Human Genetics [Internet]. 2024 [cited 2024 Aug 26]; 10.1016/j.ajhg.2024.07.018

43. Fawzy M, Marsh JA. Understanding the heterogeneous performance of variant effect predictors across human protein-coding genes. Sci Rep. Nature Publishing Group; 2024;14:26114. 10.1038/s41598-024-76202-6

44. van Loggerenberg W, Sowlati-Hashjin S, Weile J, Hamilton R, Chawla A, Sheykhkarimli D, et al. Systematically testing human HMBS missense variants to reveal mechanism and pathogenic variation. Am J Hum Genet. 2023;110:1769–86. 10.1016/j.ajhg.2023.08.012

45. Gebbia M, Zimmerman D, Jiang R, Nguyen M, Weile J, Li R, et al. A missense variant effect map for the human tumor-suppressor protein CHK2. The American Journal of Human Genetics. Elsevier; 2024;111:2675–92. 10.1016/j.ajhg.2024.10.013

46. Wright CF, Campbell P, Eberhardt RY, Aitken S, Perrett D, Brent S, et al. Genomic Diagnosis of Rare Pediatric Disease in the United Kingdom and Ireland. New England Journal of Medicine. Massachusetts Medical Society; 2023;388:1559–71. 10.1056/NEJMoa2209046

47. Stolyarova A, Coop G, Przeworski M. The distribution of highly deleterious variants across human ancestry groups. Proc Natl Acad Sci U S A. 2025;122:e2503857122. 10.1073/pnas.2503857122

48. Tavtigian SV, Harrison SM, Boucher KM, Biesecker LG. Fitting a naturally scaled point system to the ACMG/AMP variant classification guidelines. Hum Mutat. 2020;41:1734–7. 10.1002/humu.24088

49. Garrett A, Durkie M, Callaway A, Burghel GJ, Robinson R, Drummond J, et al. Combining evidence for and against pathogenicity for variants in cancer susceptibility genes: CanVIG-UK consensus recommendations. Journal of Medical Genetics. BMJ Publishing Group Ltd; 2021;58:297–304. 10.1136/jmedgenet-2020-107248

50. Guidugli L, Rugani C, Lombardi G, Aretini P, Galli A, Caligo MA. A recombination-based method to characterize human BRCA1 missense variants. Breast Cancer Res Treat. 2011;125:265–72. 10.1007/s10549-010-1112-8

51. Caligo MA, Bonatti F, Guidugli L, Aretini P, Galli A. A yeast recombination assay to characterize human BRCA1 missense variants of unknown pathological significance. Human Mutation. 2009;30:123–33. 10.1002/humu.20817

52. Cecco LD, Melissari E, Mariotti V, Iofrida C, Galli A, Guidugli L, et al. Characterisation of gene expression profiles of yeast cells expressing BRCA1 missense variants. European Journal of Cancer. Elsevier; 2009;45:2187–96. 10.1016/j.ejca.2009.04.025

53. Iofrida C, Melissari E, Mariotti V, Guglielmi C, Guidugli L, Caligo MA, et al. Effects on human transcriptome of mutated BRCA1 BRCT domain: A microarray study. BMC Cancer [Internet]. Springer Science and Business Media LLC; 2012 [cited 2025 Jul 26];12. 10.1186/1471-2407-12-207

54. Bouwman P, van der Gulden H, van der Heijden I, Drost R, Klijn CN, Prasetyanti P, et al. A High-Throughput Functional Complementation Assay for Classification of BRCA1 Missense Variants. Cancer Discovery. 2013;3:1142–55. 10.1158/2159-8290.CD-13-0094

55. Mukherjee S, Feig M. Conformational Change in MSH2-MSH6 upon Binding DNA Coupled to ATPase Activity. Biophysical Journal. Elsevier; 2009;96:L63–5. 10.1016/j.bpj.2009.04.012

56. Ollodart AR, Yeh C-LC, Miller AW, Shirts BH, Gordon AS, Dunham MJ. Multiplexing mutation rate assessment: determining pathogenicity of Msh2 variants in Saccharomyces cerevisiae. Genetics. 2021;218:iyab058. 10.1093/genetics/iyab058

57. Ko J-L, Chiao M-C, Chang S-L, Lin P, Lin J-C, Sheu G-T, et al. A novel p53 mutant retained functional activity in lung carcinomas. DNA Repair (Amst). 2002;1:755–62. 10.1016/s1568-7864(02)00094-0

58. Kato S, Han S-Y, Liu W, Otsuka K, Shibata H, Kanamaru R, et al. Understanding the function-structure and function-mutation relationships of p53 tumor suppressor protein by high-resolution missense mutation analysis. Proc Natl Acad Sci U S A. 2003;100:8424–9. 10.1073/pnas.1431692100

59. Tung M-C, Lin P-L, Wang Y-C, He T-Y, Lee M-C, Yeh SD, et al. Mutant p53 confers chemoresistance in non-small cell lung cancer by upregulating Nrf2. Oncotarget. 2015;6:41692–705. 10.18632/oncotarget.6150

60. Livesey BJ, Marsh JA. Why variant effect predictors and multiplexed assays agree and disagree [Internet]. bioRxiv; 2025 [cited 2025 Aug 1]. p. 2025.07.31.667868. 10.1101/2025.07.31.667868

61. Adamovich AI, Diabate M, Banerjee T, Nagy G, Smith N, Duncan K, et al. The functional impact of BRCA1 BRCT domain variants using multiplexed DNA double-strand break repair assays. The American Journal of Human Genetics. 2022;109:618–30. 10.1016/j.ajhg.2022.01.019

62. Jaganathan K, Kyriazopoulou Panagiotopoulou S, McRae JF, Darbandi SF, Knowles D, Li YI, et al. Predicting Splicing from Primary Sequence with Deep Learning. Cell. 2019;176:535–548.e24. 10.1016/j.cell.2018.12.015

63. Tadaka S, Katsuoka F, Ueki M, Kojima K, Makino S, Saito S, et al. 3.5KJPNv2: an allele frequency panel of 3552 Japanese individuals including the X chromosome. Hum Genome Var. 2019;6:28. 10.1038/s41439-019-0059-5

64. Gripp KW, Sol-Church K, Smpokou P, Graham GE, Stevenson DA, Hanson H, et al. An attenuated phenotype of Costello syndrome in three unrelated individuals with a HRAS c.179G>A (p.Gly60Asp) mutation correlates with uncommon functional consequences. Am J Med Genet A. 2015;167A:2085–97. 10.1002/ajmg.a.37128

65. Gripp KW, Kolbe V, Brandenstein LI, Rosenberger G. Attenuated phenotype of Costello syndrome and early death in a patient with an HRAS mutation (c.179G>T; p.Gly60Val) affecting signalling dynamics. Clin Genet. 2017;92:332–7. 10.1111/cge.12980

66. Warren JJ, Pohlhaus TJ, Changela A, Iyer RR, Modrich PL, Beese LS. Structure of the Human MutSα DNA Lesion Recognition Complex. Molecular Cell. Elsevier; 2007;26:579–92. 10.1016/j.molcel.2007.04.018

67. de Oliveira JM, Zurro NB, Coelho AVC, Caraciolo MP, de Alexandre RB, Cervato MC, et al. The genetics of hereditary cancer risk syndromes in Brazil: a comprehensive analysis of 1682 patients. Eur J Hum Genet. Nature Publishing Group; 2022;30:818–23. 10.1038/s41431-022-01098-7

68. Guindalini RSC, Viana DV, Kitajima JPFW, Rocha VM, López RVM, Zheng Y, et al. Detection of germline variants in Brazilian breast cancer patients using multigene panel testing. Sci Rep. Nature Publishing Group; 2022;12:4190. 10.1038/s41598-022-07383-1

69. Matreyek KA, Starita LM, Stephany JJ, Martin B, Chiasson MA, Gray VE, et al. Multiplex assessment of protein variant abundance by massively parallel sequencing. Nat Genet. 2018;50:874–82. 10.1038/s41588-018-0122-z

70. Tan M-H, Mester J, Peterson C, Yang Y, Chen J-L, Rybicki LA, et al. A clinical scoring system for selection of patients for PTEN mutation testing is proposed on the basis of a prospective study of 3042 probands. Am J Hum Genet. 2011;88:42–56. 10.1016/j.ajhg.2010.11.013

71. Nizialek EA, Mester JL, Dhiman VK, Smiraglia DJ, Eng C. KLLN epigenotype-phenotype associations in Cowden syndrome. Eur J Hum Genet. 2015;23:1538–43. 10.1038/ejhg.2015.8

72. Momozawa Y, Iwasaki Y, Parsons MT, Kamatani Y, Takahashi A, Tamura C, et al. Germline pathogenic variants of 11 breast cancer genes in 7,051 Japanese patients and 11,241 controls. Nat Commun. 2018;9:4083. 10.1038/s41467-018-06581-8

73. Heinrich F, Chakravarthy S, Nanda H, Papa A, Pandolfi PP, Ross AH, et al. The PTEN Tumor Suppressor Forms Homodimers in Solution. Structure. 2015;23:1952–7. 10.1016/j.str.2015.07.012

74. Thukral SK, Lu Y, Blain GC, Harvey TS, Jacobsen VL. Discrimination of DNA binding sites by mutant p53 proteins. Mol Cell Biol. 1995;15:5196–202. 10.1128/MCB.15.9.5196

75. Saller E, Tom E, Brunori M, Otter M, Estreicher A, Mack DH, et al. Increased apoptosis induction by 121F mutant p53. EMBO J. 1999;18:4424–37. 10.1093/emboj/18.16.4424

76. Kaeser MD, Iggo RD. Chromatin immunoprecipitation analysis fails to support the latency model for regulation of p53 DNA binding activity in vivo. Proc Natl Acad Sci U S A. 2002;99:95–100. 10.1073/pnas.012283399

77. Kakudo Y, Shibata H, Otsuka K, Kato S, Ishioka C. Lack of correlation between p53-dependent transcriptional activity and the ability to induce apoptosis among 179 mutant p53s. Cancer Res. 2005;65:2108–14. 10.1158/0008-5472.CAN-04-2935

78. Inga A, Resnick MA. Novel human p53 mutations that are toxic to yeast can enhance transactivation of specific promoters and reactivate tumor p53 mutants. Oncogene. 2001;20:3409–19. 10.1038/sj.onc.1204457

79. Desmond A, Kurian AW, Gabree M, Mills MA, Anderson MJ, Kobayashi Y, et al. Clinical Actionability of Multigene Panel Testing for Hereditary Breast and Ovarian Cancer Risk Assessment. JAMA Oncol. 2015;1:943–51. 10.1001/jamaoncol.2015.2690

80. Sondka Z, Dhir NB, Carvalho-Silva D, Jupe S, Madhumita, McLaren K, et al. COSMIC: a curated database of somatic variants and clinical data for cancer. Nucleic Acids Research. 2024;52:D1210– 7. 10.1093/nar/gkad986

81. Fortuno C, Cipponi A, Ballinger ML, Tavtigian SV, Olivier M, Ruparel V, et al. A quantitative model to predict pathogenicity of missense variants in the TP53 gene. Hum Mutat. 2019;40:788–800. 10.1002/humu.23739

82. Karczewski KJ, Francioli LC, Tiao G, Cummings BB, Alföldi J, Wang Q, et al. The mutational constraint spectrum quantified from variation in 141,456 humans. Nature. Nature Publishing Group; 2020;581:434–43. 10.1038/s41586-020-2308-7

83. Danecek P, Bonfield JK, Liddle J, Marshall J, Ohan V, Pollard MO, et al. Twelve years of SAMtools and BCFtools. GigaScience. 2021;10:giab008. 10.1093/gigascience/giab008

84. McLaren W, Gil L, Hunt SE, Riat HS, Ritchie GRS, Thormann A, et al. The Ensembl Variant Effect Predictor. Genome Biology. 2016;17:122. 10.1186/s13059-016-0974-4

85. Berman HM, Westbrook J, Feng Z, Gilliland G, Bhat TN, Weissig H, et al. The Protein Data Bank. Nucleic Acids Res. 2000;28:235–42.

86. Agirre J, Atanasova M, Bagdonas H, Ballard CB, Baslé A, Beilsten-Edmands J, et al. The CCP4 suite: integrative software for macromolecular crystallography. Acta Cryst D. International Union of Crystallography; 2023;79:449–61. 10.1107/S2059798323003595

87. Miller S, Janin J, Lesk AM, Chothia C. Interior and surface of monomeric proteins. Journal of Molecular Biology. Academic Press; 1987;196:641–56. 10.1016/0022-2836(87)90038-6

88. Levy ED. A Simple Definition of Structural Regions in Proteins and Its Use in Analyzing Interface Evolution. Journal of Molecular Biology. Academic Press; 2010;403:660–70. 10.1016/j.jmb.2010.09.028

89. Delgado J, Radusky LG, Cianferoni D, Serrano L. FoldX 5.0: working with RNA, small molecules and a new graphical interface. Bioinformatics. 2019;35:4168–9. 10.1093/bioinformatics/btz184

