## Supplementary figures and images for "Combining MAVEs and computational predictors improves variant classification across ancestries in hereditary cancer genes"

### FigS1.jpeg

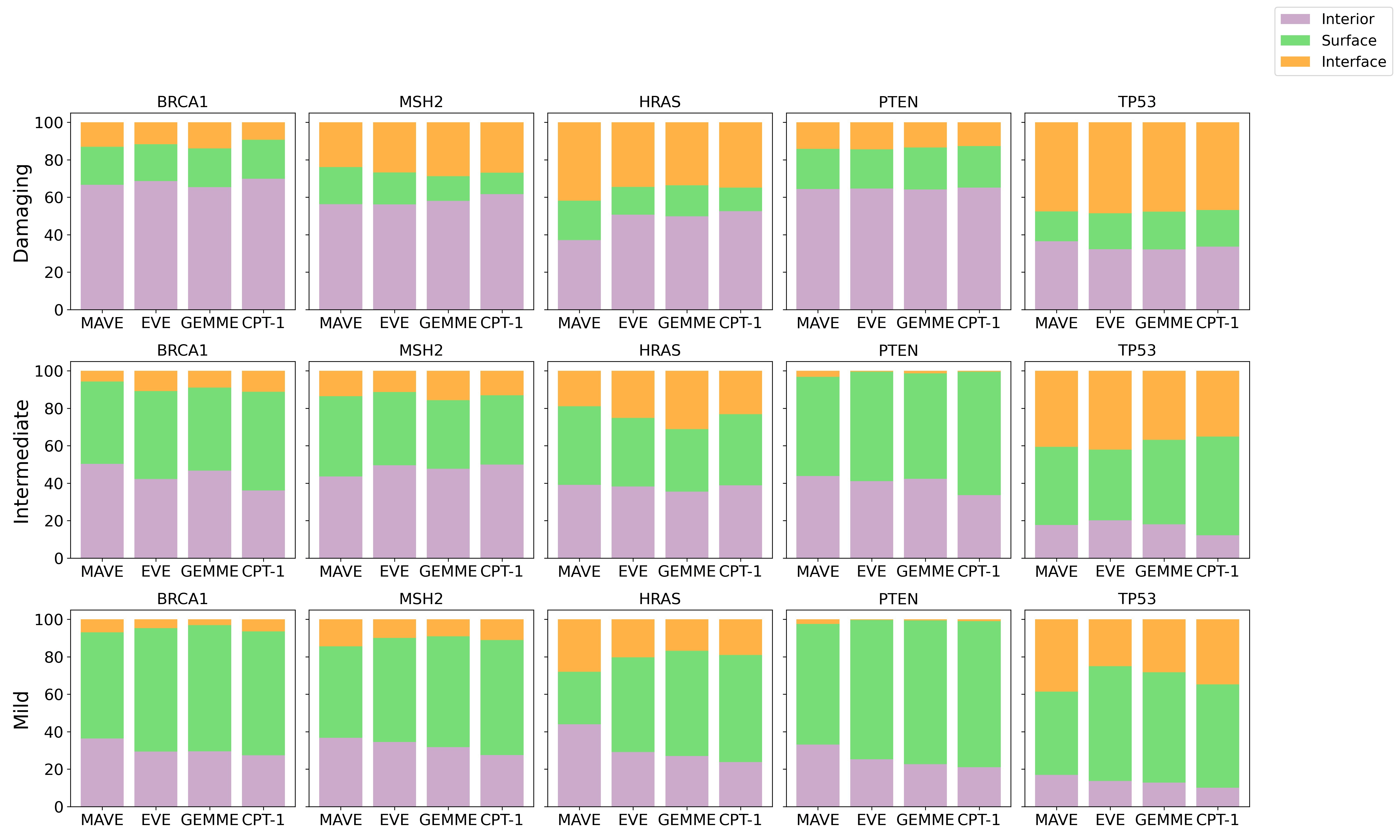

### FigS2.jpeg

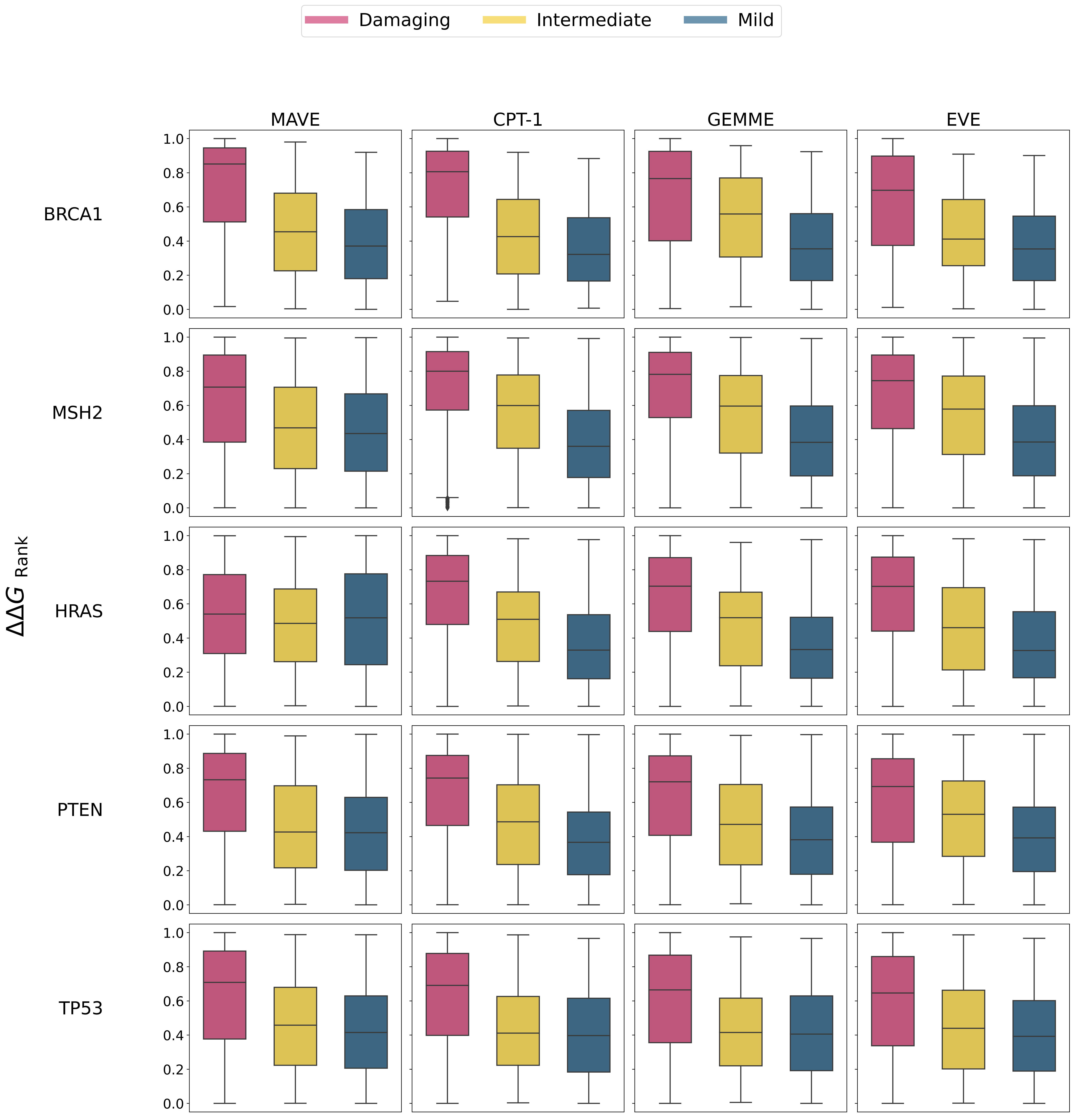

### FigS3.jpeg

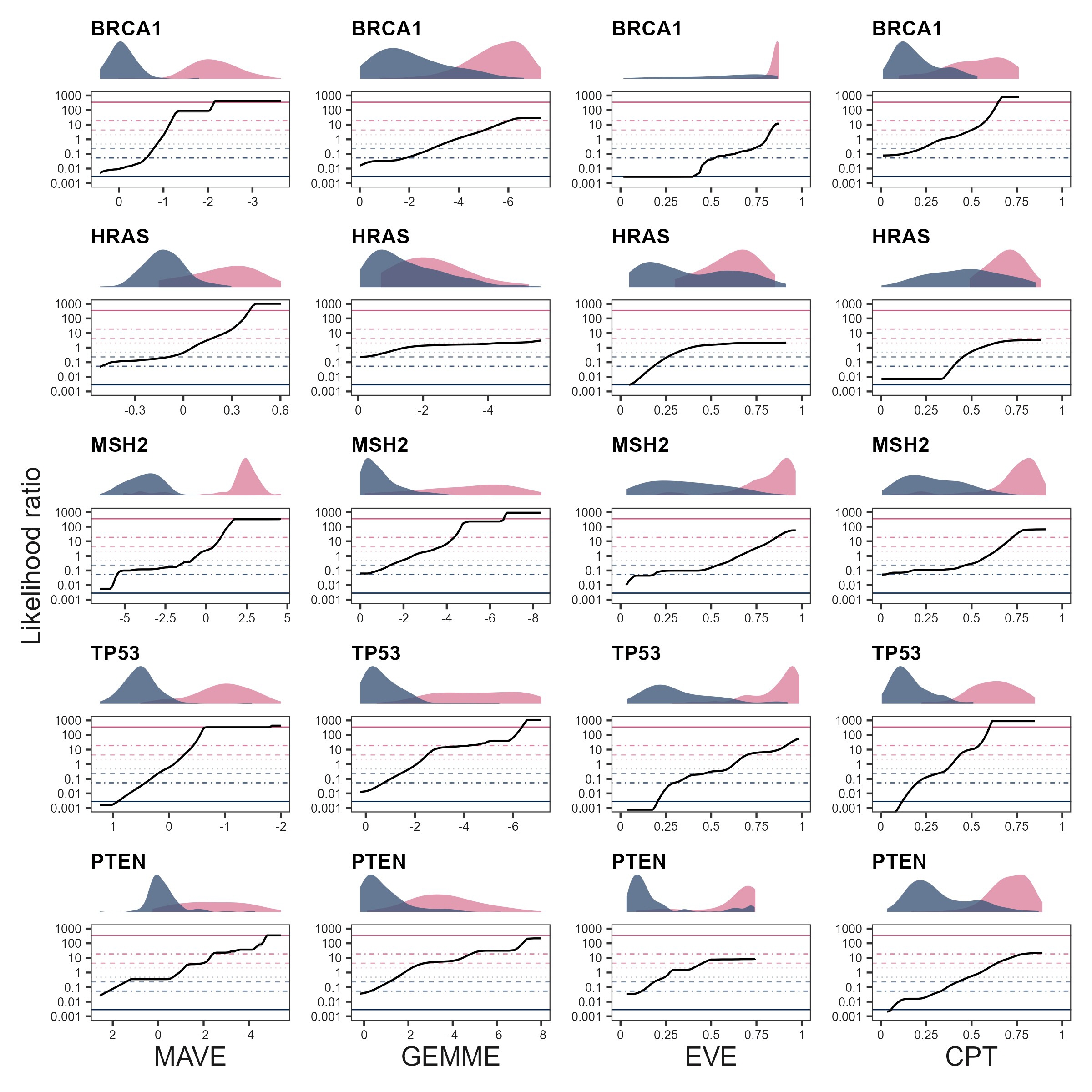
